# Development, External Validation, and Biomolecular Corroboration of Interoperable Models for Identifying Critically Ill Children at Risk of Neurologic Morbidity

**DOI:** 10.1101/2024.09.17.24313649

**Authors:** Christopher M. Horvat, Amie J Barda, Eddie Perez Claudio, Alicia K. Au, Andrew Bauman, Qingyan Li, Ruoting Li, Neil Munjal, Mark Wainwright, Tanupat Boonchalermvichien, Harry Hochheiser, Robert S. B. Clark

**Affiliations:** Department of Critical Care Medicine, University of Pittsburgh, Pittsburgh, PA; Safar Center for Resuscitation Research, University of Pittsburgh, Pittsburgh, PA; Barda Analytics Consulting LLC, North Royalton, OH; Department of Biomedical Informatics, University of Pittsburgh, Pittsburgh, PA; Seattle Children’s Hospital, Seattle, WA; Department of Pediatrics, University of Wisconsin-Madison, Madison, WI

## Abstract

**Importance:** Declining mortality in the field of pediatric critical care medicine has shifted practicing clinicians’ attention to preserving patients’ neurodevelopmental potential as a main objective. Earlier identification of critically ill children at risk for incurring neurologic morbidity would facilitate heightened surveillance that could lead to timelier clinical detection, earlier interventions, and preserved neurodevelopmental trajectory.

**Objective:** Develop machine-learning models for identifying acquired neurologic morbidity while hospitalized with critical illness and assess correlation with contemporary serum-based, brain injury-derived biomarkers.

**Design:** Retrospective cohort study.

**Setting:** Two large, quaternary children’s hospitals.

**Exposures:** Critical illness.

**Main Outcomes and Measures:** The outcome was neurologic morbidity, defined according to a computable, composite definition at the development site or an order for neurocritical care consultation at the validation site. Models were developed using varying time windows for temporal feature engineering and varying censored time horizons prior to identified neurologic morbidity. Optimal models were selected based on F1 scores, cohort sizes, calibration, and data availability for eventual deployment. A generalizable created at the development site was assessed at an external validation site and optimized with spline recalibration. Correlation was assessed between development site model predictions and measurements of brain biomarkers from a convenience cohort.

**Results:** After exclusions there were 14,222-25,171 encounters from 2010-2022 in the development site cohorts and 6,280-6,373 from 2018-2021 in the validation site cohort. At the development site, an extreme gradient boosted model (XGBoost) with a 12-hour time horizon and 48-hour feature engineering window had an F1-score of 0.54, area under the receiver operating characteristics curve (AUROC) of 0.82, and a number needed to alert (NNA) of 2. A generalizable XGBoost model with a 24-hour time horizon and 48-hour feature engineering window demonstrated an F1-score of 0.37, AUROC of 0.81, AUPRC of 0.51, and NNA of 4 at the validation site. After recalibration at the validation site, the Brier score was 0.04. Serum levels of the brain injury biomarker glial fibrillary acidic protein measurements significantly correlated with model output (r_s_=0.34; *P*=0.007).

**Conclusions and Relevance:** We demonstrate a well-performing ensemble of models for predicting neurologic morbidity in children with biomolecular corroboration. Prospective assessment and refinement of biomarker-coupled risk models in pediatric critical illness is warranted.

**Key Points:** **Question** Can interoperable models for predicting neurological deterioration in critically ill children be developed, correlated with serum-based brain-derived biomarkers, and validated at an external site?

**Findings** A development site model demonstrated an area under the receiver operating characteristics curve (AUROC) of 0.82 and a number needed to alert (NNA) of 2. Predictions correlated with levels of glial fibrillary acidic protein in a subset of children. A generalizable model demonstrated an AUROC of 0.81 and NNA of 4 at the validation site.

**Meaning** Well performing prediction models coupled with brain biomarkers may help to identify critically ill children at risk for acquired neurological morbidity.

## Introduction

An estimated 340,000 children are hospitalized with critical illness every year in the United States and brain injury has been cited as the proximate cause of death in approximately 90% of previously healthy children who do not survive their intensive care admission.^1,2^ Of children who survive critical illness, acquired neurologic morbidity can have long-lasting implications which range from mild impairments in cognition to profound debilitation. Declining mortality in the field of pediatric critical care has led to increased attention to the longer-term functional outcomes of children who survive an intensive care admission.^3^

Granular, time-series data harbored by the electronic health record (EHR) offer a rich training ground for probabilistic models of important patient outcomes. Implementing well-performing models as clinical decision support (CDS) systems is a promising approach for improving outcomes related to many different conditions and situations, though there are currently no established tools for identifying children at risk for new brain injury.^4–7^ Recently enacted federal mandates in the United States of America (USA) are promoting the development of EHRs that facilitate the deployment of interoperable decision support tools built to leverage a core dataset.^8^

The main objective of the present work was to construct and externally validate predictive models to support the identification of critically ill children at high risk for acquired neurologic morbidity, as a first step towards the development of a decision support tool that might be used to forewarn of neurologic morbidity amongst critically ill children, as well as to aid in the enrichment of prospective trials examining strategies to mitigate the risk of brain injury during pediatric critical illness. A second objective was to corroborate the biological underpinnings of the developed prediction models by assessing correlation with novel, brain-derived, serum-based biomarkers of brain injury obtained from a diagnostically diverse cohort of critically ill children.

## Methods

### Study Sites

Model development used data from all encounters to a quaternary pediatric intensive care unit (PICU) in a large, freestanding children’s hospital between January 1, 2010 and December 31, 2022. The development site PICU serves a region of approximately 5 million people, encompassing Western Pennsylvania and bordering states, and is a level 1 pediatric trauma center. External model validation occurred using data from encounters admitted between January 1, 2018 and December 31, 2023 to a quaternary PICU in a large, freestanding children’s hospital that serves as a referral center for the 5-state region of Washington, Wyoming, Alaska, Montana, and Idaho. Approval was granted by the institutional review boards of the University of Pittsburgh (Institutional Review Board [IRB] #17030743) and Seattle Children’s Hospital (IRB #STUDY00001374). Findings are reported according to the Transparent Reporting of a Multivariable Prediction Model for Individual Prognosis or Diagnosis (TRIPOD) statement (**Supplemental Table 1**).

### Model Development Frameworks

Conceptualization of the model adhered to the Littenberg framework for the development of clinical decision support tools, which considers the clinical and technical plausibility of the tool, as well as the process outcomes, patient outcomes, and eventual societal outcomes addressed by the tool (**Supplemental Table 2**).^9^ The first 5 steps of the cross-industry standard process for data modeling (CRISP-DM) framework were followed for model design. CRISP-DM outlines six steps for data science projects that include 1) understanding the use case; 2) understanding the data; 3) data curation; 4) model development; 5) model evaluation; and 6) model deployment.^10^

### Model Development Approach

Model development proceeded in 2 phases: 1) Development of models for use locally at the development site; 2) Development of generalizable models with external validation. The outcome of neurologic morbidity was defined using structured EHR data surrogates based on each study site’s clinical and electronic workflows. At the development site, the outcome was a previously validated, computable, composite definition of neurologic morbidity that incorporated orders for electroencephalography (EEG), brain computed tomography (CT), brain magnetic resonance imaging (MRI), or indicators of treated delirium within 72-hours of one another (**Supplemental Table 3**).^11^ This outcome has also been validated in a separate cohort of children with sepsis.^12^ At the validation site, orders for a neurocritical care service consultation were deemed to be the most reliable surrogate for neurologic morbidity during an episode of critical illness. Data for control cases (hospitalized children who did not meet the definition of a neurologic morbidity) were collected from a random period during the encounter with preference to a window following the first PICU admission.

Candidate data elements for model construction were selected based on clinical expertise and with attention to the United States Core Data for Interoperability (USCDI) requirements to facilitate eventual, interoperable deployment (**Supplemental Tables 4 and 5**).^13^ A ‘Biodigital Rapid Alert to Identify Neurologic morbidity, A-I bundle (BRAIN A-I)’ standard clinical vocabulary value set was filed with the National Library of Medicine’s Value Set Authority Center.^14^ Features were engineered with the dual aims of representing the temporality of the data while also preserving clinical interpretability of the features, using methods previously reported.^15^ Features were then discretized, or categorized into information bins, with missingness encoded as a feature. In addition to preserving possible information associated with missingness, discretization was performed to reduce the influence of outlier data, represent data nonlinearity in linear modeling processes such as logistic regression, further mitigate overfitting, and preserve clinical interpretability of the features. Additional details of data curation and model development are in the **Supplemental Model Methods**. **Supplemental Figure 4** summarizes model construction at the development site and evaluation at the external validation site. At the development site, data were queried from an Oracle (Oracle Corp, Austin, TX) data warehouse containing a subset of transformed tables from the Cerner Millennium database (Oracle Cerner, Kansas City, MO). The model was developed and assessed using Python (version [v]3.10.11), Jupyter (v1.0.0), and the packages *Pandas* (v1.5.3), *Numpy* (v1.25.0), *Matplotlib* (v3.71.1), *Sklearn* (v1.1.1), *XGBoost* (v1.7.3), Seaborn (v0.11.2), SHAP (v0.41.0), and *tqdm* (v4.65.0).

### Biomolecular Corroboration of the Model at the Development Site

Model predictions were compared to measured levels of 6 serum-based, brain-derived biomarkers of brain injury obtained from a previously assembled convenience cohort of 101 children hospitalized between 2012-2014 (IRB #19040172). The biomarkers were ubiquitin C-terminal hydrolase-L1 (UCH-L1), glial fibrillary acidic protein (GFAP), myelin basic protein (MBP), neuron-specific enolase (NSE), S100 calcium binding protein B (S100B), and spectrin breakdown product 150 (SBDP150). After prospective consent from a legal guardian, biomarker levels were collected for up to 7 consecutive days from critically ill children with preexisting central venous catheters or arterial catheters. Details of the assays are provided in the **Supplemental Biomarker Methods**. Maximum values of each biomarker for each encounter were assessed for correlation with the predicted probability of neurologic deterioration for that encounter. Patients were determined to have a neurologic complication by chart review if it occurred no more than 7 days after the last date a biomarker was collected.

### Model Selection and Statistical Analysis

The top-performing model was selected based on F1 score, considering a clinically actionable time horizon, as well as the volume of available training data for feature engineering. Models with <0.15 difference in F1 scores were then compared both by visual inspection of calibration plots and Brier scores. Additional Fβ thresholds of 0.5, 2, and 3 were secondarily evaluated to identify whether there were any substantial differences in the optimal classifier based on the relative weight of recall compared to precision. Statistical performances of the top-performing models were evaluated at varied model outputs ranging from 0.025 to 0.9. Spline regression was performed on top-performing models to improve calibration. Normally distributed continuous data are presented as means and 95% confidence intervals, nonparametric continuous data are presented as medians with interquartile ranges (IQRs), and categorical data are presented as counts with corresponding proportions. Model discrimination was compared to the discrimination of the last Glasgow coma scale (GCS) score prior to the censored time horizon using DeLong’s method. For the biomolecular corroboration analysis, Spearman’s rank-order correlation was assessed between a chart-adjudicated neurologic morbidity outcome and the composite neurologic morbidity outcome, as well as between the probability output of top-performing models and the composite neurologic morbidity outcome. Correlation was then assessed between biomarker levels and the probability output of the top-performing models.

Notched boxplots with overlying violin plots were constructed for significantly correlated biomarkers by dichotomizing predicted neurologic morbidity according to whether the probability was <0.5 or ≥0.5. The distributions of biomarker measurements were normalized for plotting using log transformation and significance testing was assessed using an independent *t* test. An α < 0.05 is considered significant. Statistical analyses not performed in Python were performed in R version 4.3.1 (R Foundation, Vienna, Austria).

## Results

### Development Site Models Performance

There were 32,702 encounters with a PICU stay. After exclusions, cohort sizes ranged from 14,222-25,171 encounters, with 18,568 encounters in the final model cohort (**Supplemental Table 8; Figure 1A**). Patients were slightly older, received less mechanical ventilation, and less sedative-analgesic medications in the final test dataset compared to the training and validation datasets (**Table 1**). The final models evaluated in the test dataset was the extreme gradient boosting (XGBoost) model with a 12-hour time horizon and 48-hour feature window. This model was determined by investigator agreement to be a reasonable balance of favorable F1 scores, calibration as assessed by a Brier score, visual inspection of the calibration plot, clinically actionable time horizon, and sufficient cohort size for the training, validation, and test datasets.

**Figure 1.**
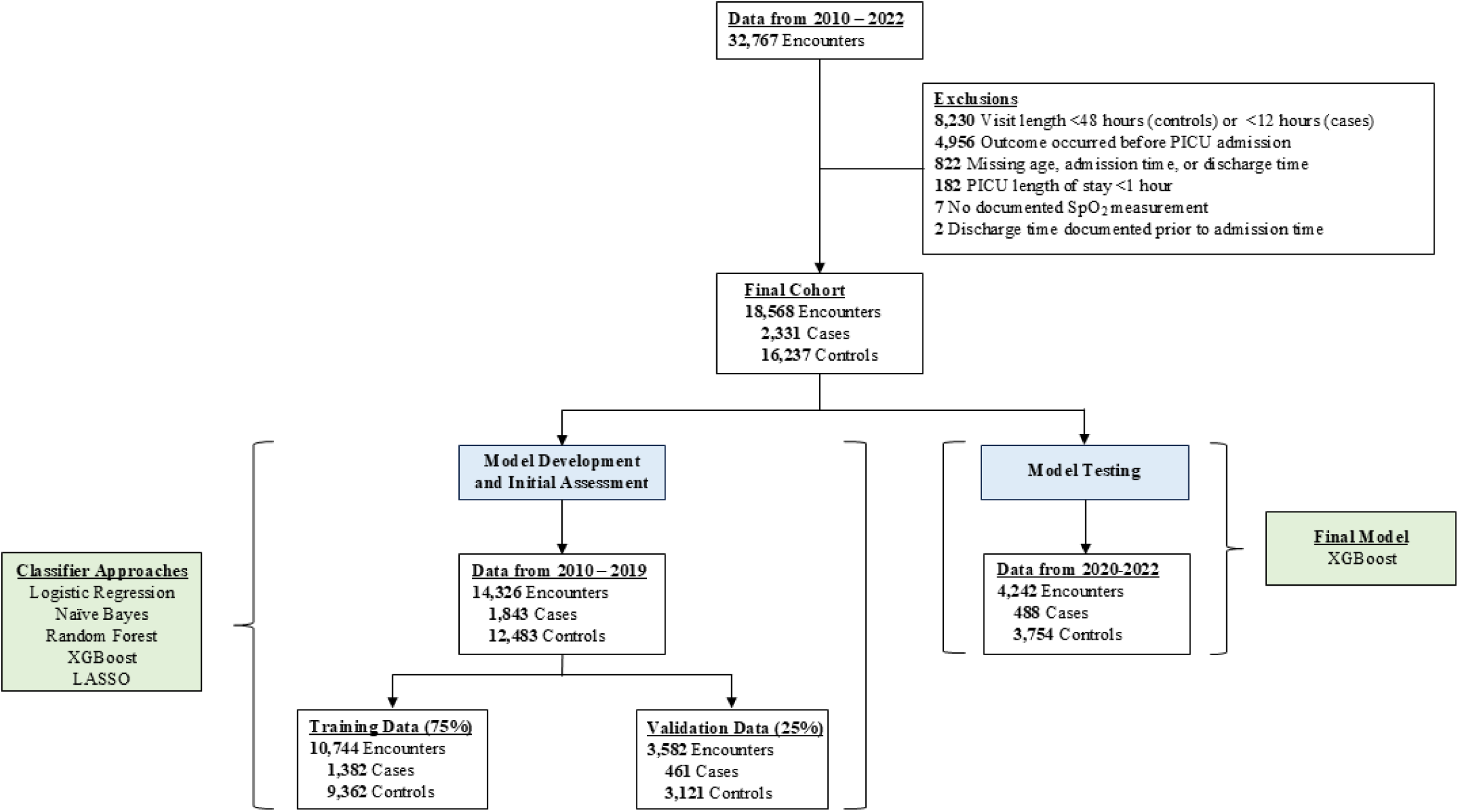
Cohort ascertainment for the final model at the development site, which included features engineered using 48 hours of preceding data and censoring 12-hours prior to the event for cases. Initial model development and validation proceeded using data from 2010-2019. The model was tested using data from 2020-2022. Abbreviations: LASSO, least absolute shrinkage and selection operator; PICU, pediatric intensive care unit; XGBoost, extreme gradient boosting.

**Table 1.** Demographic characteristics of the entire cohort and the parsed training, validation, and final test datasets at the development site and the entire cohort, training, and validation datasets, as well as the generalizable model validation dataset at the external validation site.

| Characteristic | Development Site |  |  |  | Validation Site |
| --- | --- | --- | --- | --- | --- |
|  | Entire Cohort | Training Dataset | Validation Dataset | Final Test Dataset | Generalizable Model Validation Dataset |
|  | **N = 18,568** | **n = 10,744** | **n = 3,582** | **n = 4,242** | **N = 6,825** |
| Age (months), median (IQR) | 70 (18, 161) | 67 (18, 158) | 68.5 (18, 160) | 77 (19, 167) | 96 (18,171) |
| Female, n (%) | 8,325 (45) | 4,748 (44) | 1,593 (44) | 1,984 (47) | 3,159 (46) |
| Glasgow Coma Scale Score, median (IQR)* | 15 (12, 15) | 15 (11, 15) | 15 (11, 15) | 15 (14, 15) | 14 (14,15) |
| Mechanical Ventilation, n (%) | 5,352 (29) | 3,322 (31) | 1,143 (32) | 887 (21) | 1,948(29) |
| Endotracheal Tube, n (%) | 1,759 (9) | 1,142 (11) | 370 (10) | 247 (6) | 1,283 (19) |
| Vasoactive Medication, n (%)** | 720 (4) | 393 (4) | 148 (4) | 179 (4) | 221 (3) |
| Sedative-Analgesic Medication, n (%)*** | 4,660 (25) | 2,948 (27) | 987 (27) | 727 (17) | 1,930 (28) |
| *The last recorded Glasgow coma scale score for the encounter prior to the censored time horizon<br>**Vasoactive medications include dobutamine, dopamine, epinephrine, norepinephrine, or milrinone<br>***Sedative-analgesic medications include fentanyl, hydromorphone, midazolam, or morphine<br>Abbreviations: IQR, interquartile range |  |  |  |  |  |

Complete development site model performance characteristics, 1 for each of the combinations of a 6, 12, and 24-hour censored time horizons and 24, 48, and 72-hour feature windows selected based on F1 score, are detailed in **Supplemental Table 9**. Each approach generated 605 features prior to information gain feature selection. The F1 scores are reported in **Supplemental Tables 10A and 10B**. Additional Fβ scores largely agreed with the model assessments provided by F1 scores and are presented in **Supplemental Table 11A-C**.

The final model contained 352 features and had a number needed to alert (NNA) of 2 when considering a model prediction of greater than or equal to 0.5 as positive. At a model prediction threshold of 0.025 in the test dataset, sensitivity increased to 0.86 and NAA was 4. Statistical performance of the top-performing validation models and final test model at a range of output thresholds are in **Supplemental Table 12A and 12B** and **Supplemental Figure 5**. All development site models had a NNA of 2-3 at this prediction threshold. In the test dataset the final model had a sensitivity of 0.47 (range for all models in the validation dataset [range] 0.24-0.63), specificity of 0.98 (range 0.96-0.99), AUPRC 0.68 (range 0.39-0.78), and AUROC of 0.89 (range 0.80-0.87). The final model had significantly greater discrimination compared to the last GCS AUROC of 0.72 obtained prior to the censored time horizon, *P*<0.001. Calibration plots of models with comparable performance based on F1-scores are displayed in **Supplemental Figure 6**. The top 10 features of the final model are displayed in **Supplemental Figure 7**. Average hourly scores for cases and controls for the 12 hours preceding and 4 hours following an outcome event are displayed in **Figure 2**.

**Figure 2.**
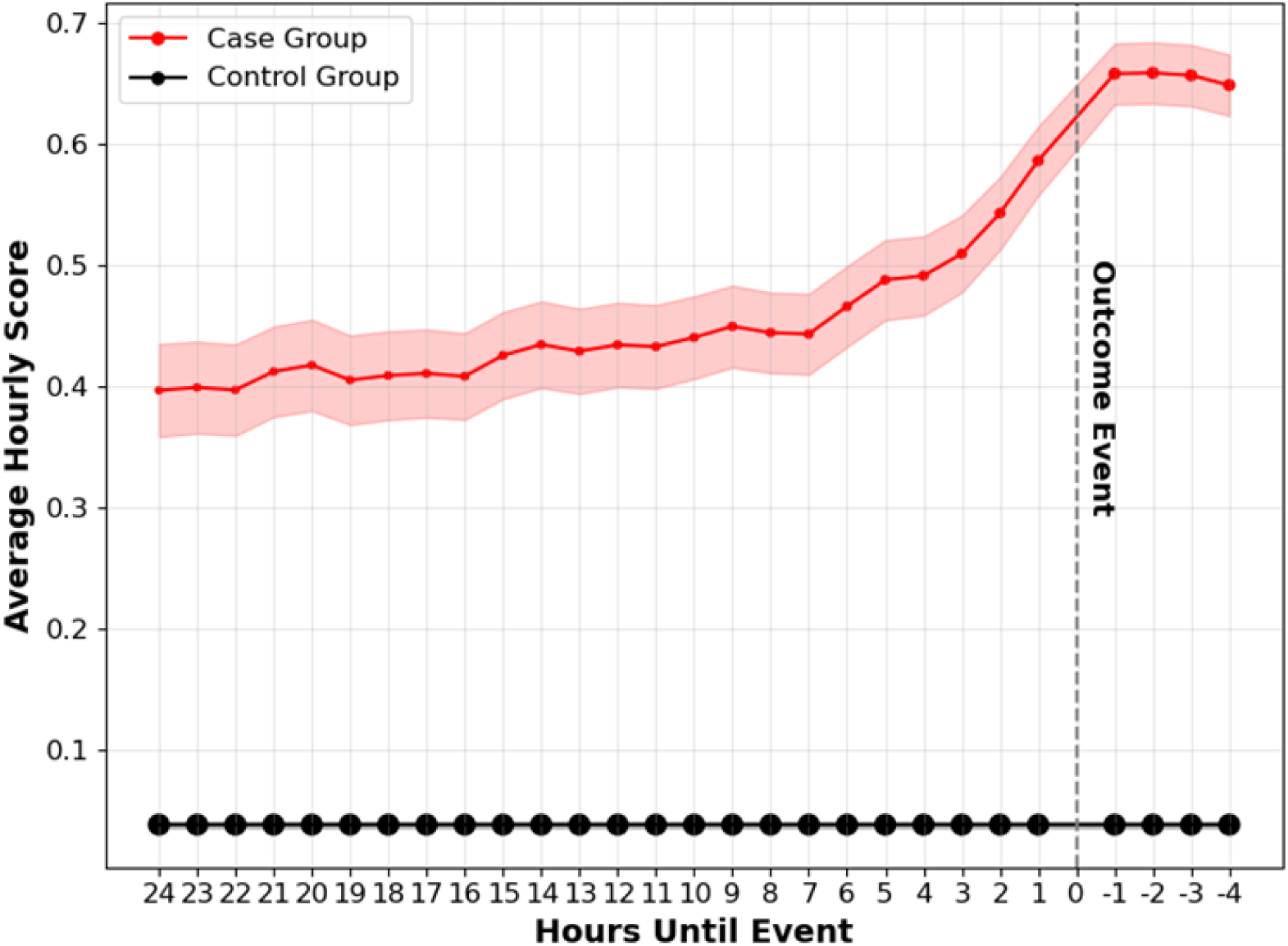
Average hourly scores in the test dataset (encounters with a PICU stay in the year 2020 – 2022) for varied censored time horizon windows for the extreme gradient boosted model developed using a 12-hour time horizon and 48-hour feature window. The red dots are the average hourly scores 12 hours prior to an event and 4 hours after an event for the case encounters (encounters with an identified neurologic morbidity) and the shaded red region represents the 95% confidence interval. The black dots are the average hourly scores for the control encounters (encounters without an identified neurological morbidity). Confidence intervals for the control encounters are not discernable in the figure due to the large cohort size. The size of the dots is proportionate to the cohort size at that timepoint.

### Biomolecular Corroboration at the Development Site

Of the 101 patients with available brain-derived biomarkers measured, 64 also had model predictions for the 12-hour time horizon and 48-hour feature window models. Chart-adjudicated neuromorbidity within 7 days of last biomarker collection was significantly correlated with the composite neurologic morbidity outcome, r_s_=0.38 (*P*=0.002). The extreme gradient boosting model was significantly correlated with the composite neurologic morbidity outcome, r_s_=0.80 (*P*<0.001). The logistic regression model had an F1 score that was nearly identical to the extreme gradient boosting model and was also significantly correlated with the composite neurologic morbidity outcome, r_s_=0.55 (*P*<0.001). Extreme gradient boosting predictions were significantly correlated with maximum GFAP measurements, r_s_=0.34 (*P*=0.007) (**Figure 3**).

**Figure 3.**
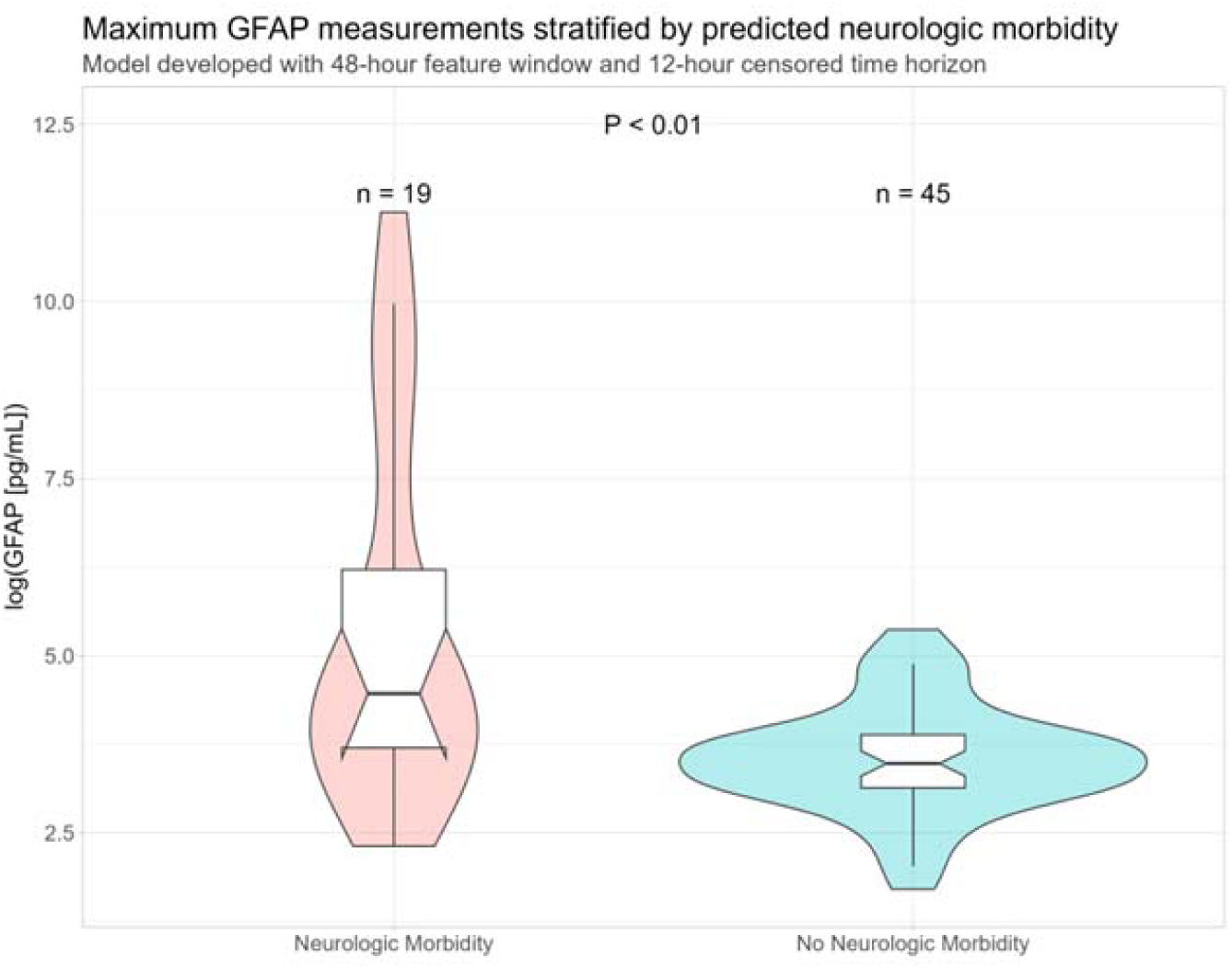
Log-transformed maximum GFAP measurements for a convenience cohort of 64 patients, stratified by predicted neurologic morbidity using the 12-hour time horizon 48-hour feature window extreme gradient boosting model. Abbreviations: GFAP, glial fibrillary acidic protein; mL, milliliter; pg, picogram.

### Generalizable Model Performance

Cohort ascertainment for the generalizable model is reported for the 24-hour time horizon and 48-hour feature window model at the development site in **Supplemental Table 8** and for the validation site in **Supplemental Table 13**. The generalizable model performance at the development and validation sites is reported in **Table 2**. Performance was comparable to earlier 24-hour time horizon 48-hour feature window models at the development. As the XGBoost and logistic regression models performed comparably, both were assessed at the validation site.

**Table 2.**
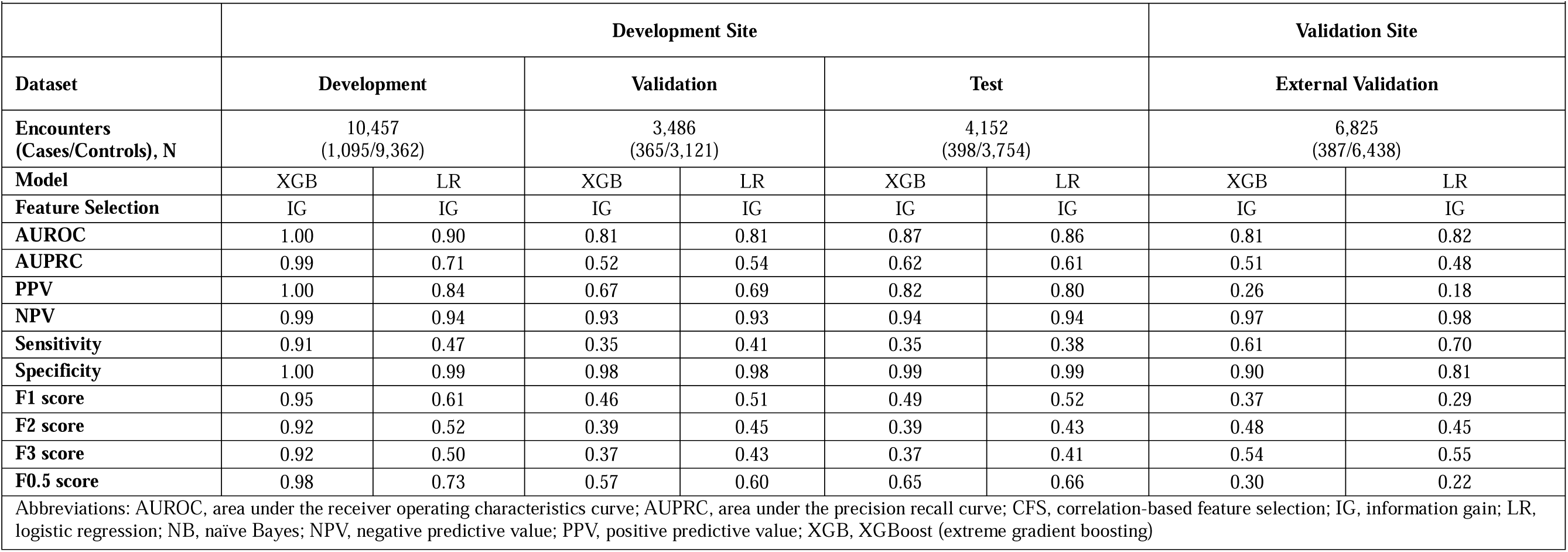
Performance of the generalizable model at the development and external validation sites.

|  | Development Site |  |  |  |  |  | Validation Site |  |
| --- | --- | --- | --- | --- | --- | --- | --- | --- |
| Dataset | Development |  | Validation |  | Test |  | External Validation |  |
| Encounters<br>(Cases/Controls), N | 10,457<br>(1,095/9,362) |  | 3,486<br>(365/3,121) |  | 4,152<br>(398/3,754) |  | 6,825<br>(387/6,438) |  |
| Model | XGB | LR | XGB | LR | XGB | LR | XGB | LR |
| Feature Selection | IG | IG | IG | IG | IG | IG | IG | IG |
| AUROC | 1.00 | 0.90 | 0.81 | 0.81 | 0.87 | 0.86 | 0.81 | 0.82 |
| AUPRC | 0.99 | 0.71 | 0.52 | 0.54 | 0.62 | 0.61 | 0.51 | 0.48 |
| PPV | 1.00 | 0.84 | 0.67 | 0.69 | 0.82 | 0.80 | 0.26 | 0.18 |
| NPV | 0.99 | 0.94 | 0.93 | 0.93 | 0.94 | 0.94 | 0.97 | 0.98 |
| Sensitivity | 0.91 | 0.47 | 0.35 | 0.41 | 0.35 | 0.38 | 0.61 | 0.70 |
| Specificity | 1.00 | 0.99 | 0.98 | 0.98 | 0.99 | 0.99 | 0.90 | 0.81 |
| F1 score | 0.95 | 0.61 | 0.46 | 0.51 | 0.49 | 0.52 | 0.37 | 0.29 |
| F2 score | 0.92 | 0.52 | 0.39 | 0.45 | 0.39 | 0.43 | 0.48 | 0.45 |
| F3 score | 0.92 | 0.50 | 0.37 | 0.43 | 0.37 | 0.41 | 0.54 | 0.55 |
| F0.5 score | 0.98 | 0.73 | 0.57 | 0.60 | 0.65 | 0.66 | 0.30 | 0.22 |
| Abbreviations: AUROC, area under the receiver operating characteristics curve; AUPRC, area under the precision recall curve; CFS, correlation-based feature selection; IG, information gain; LR, logistic regression; NB, naïve Bayes; NPV, negative predictive value; PPV, positive predictive value; XGB, XGBoost (extreme gradient boosting) |  |  |  |  |  |  |  |  |

There were 6,825 encounters in the external validation site final cohort (387 cases and 6,438 controls). As assessed by an F1 score of 0.37 at a threshold of 0.5, the top performing model was the XGBoost model, with an external validation AUROC of 0.81, AUPRC of 0.51, and an NNA of 4. Model performance characteristics across varied thresholds are displayed in **Supplemental Table 14** and **Supplemental Figure 8**. Calibration was again excellent at the development site, initially poor at the external validation site, then substantially improved after spline recalibration at the validation site (**Supplemental Figure 9**). Feature importance analysis for the generalizable model was similar between the development and validation sites (**Supplemental Figure 10**). All models outperformed the GCS, *P*<0.001.

## Discussion

In this study, we constructed well-performing models for predicting neurologic morbidity among critically ill children using EHR data from 2 large children’s hospitals. These models were trained using more than 600 features engineered to capture nonlinear relationships between predictors and the outcome. The top performing model at the development site had 352 features and a NNA of 2, suggesting the utility of incorporating more features than can be accommodated by traditional, manually-tabulated clinical decision rules. A generalizable model demonstrated robust performance at both the development site and the external validation site. All models outperformed the GCS, supporting machine-learning-based methods to facilitate clinical activities including identification of high-risk patients for clinical intervention and for identifying an enriched population for enrollment in clinical trials.^16^ By largely adhering to data elements prioritized by USCDI, the developed models have a clearer path to implementation in modern informatics architectures capable of data transfer using standard clinical vocabularies and the fast healthcare interoperability resources (FHIR) standard.^17^ The generalizable model relies on 41 variables, 37 of which are included in USCDI versions 1 or 2 and are therefore expected to ease eventual work associated with deployment.

Many predictive models are constructed utilizing a snapshot of information from a discrete moment in time.^18,19^ For predictive models to more completely leverage the content of the EHR, the temporality of data must be incorporated into model features. The performance of the present models was likely bolstered by incorporating features engineered using vector space representations of patient state, resulting in performance metrics that surpass those of other commonly used critical care risk models.^15^ The Simplified Acute Physiology Score, a commonly used mortality prediction tool for critically ill adults, has reported AUPRCs between 0.2-0.3 for in-hospital and 30-day mortality.^20^ The sequential organ failure assessment (SOFA), quick SOFA, and systemic inflammatory response syndrome criteria have reported AUPRCs of 0.06, 0.1, and 0.09 predicting mortality at the time of sepsis onset, respectively.^21^ By comparison, our model ensemble had AUPRCs ranging from 0.39-0.78 at the development site and 0.2-0.42 at the validation site. We undertook the present work with an expectation that identification of impending neurologic deterioration requires examination of contextual elements of care and more subtle vital sign and laboratory signatures which may serve as a harbinger of unfavorable trajectory.

Correlation between a top-performing model and measurements of GFAP from a convenience cohort is compatible with our previous investigations of brain biomarkers in critically ill children.^22^ GFAP is found in astrocytes and plays a role responding to central nervous system injuries and related neurodegeneration.^23^ GFAP measurements from our convenience cohort were obtained for the first 7 days of the PICU stay and may have been obtained remote from an incurred brain injury, including one detected by the composite neurologic morbidity outcome. Notably, our composite neurologic morbidity outcome was significantly correlated with chart-adjudicated neurologic morbidity and an XGBoost model was significantly associated with GFAP levels. Most extensively studied in the context of traumatic brain injury, a growing body of evidence suggest GFAP may be useful to identify more subtle insults to the central nervous system, and that the ability to measure GFAP in the bloodstream in non-traumatic diseases might relate to its dispersion into the bloodstream via recently discovered glymphatic pathways.^24,25^ Our models may prove useful both to determine for which patients a GFAP level should be obtained, as well as coupled with the GFAP measurements to bolster model performance.

This work has some important limitations. Use of a composite definition of neurological morbidity intrinsically omits occult neurological morbidities that did not trigger clinical action and represents a source of potential bias in model development. While the computable composite definition of neurologic morbidity used in the present study has previously demonstrated high specificity, the modest sensitivity of the definition suggests that the present models may miss neurologic morbidities that do not warrant inpatient imaging, EEG, a mental health assessment or a medication directed at psychosis or delirium. This limitation, however, can be mitigated by assessing performance characteristics, including varied Fβ scores or sensitivities at different output thresholds, according to context and adjusting the model actionable threshold in a manner tailored to the clinical environment in which it is deployed. Moreover, while performance was robust at an external validation site relative to other established risk scores, statistical metrics did deteriorate compared to those observed in the test dataset at the development site. Notably, the GCS also had a lower AUROC at the validation site compared to the GCS AUROC at the development site, suggesting that the choice of neurocritical care consult as an outcome influenced the models’ performance characteristics.

In conclusion, we developed well-performing models for predicting children with critical illness at risk for neurologic morbidity. A flexible, distributed strategy for model development in partnership with an external validation site demonstrated the utility of adapting to varied informatics infrastructures and EHR deployments to generate well-performing predictive models for a common clinical goal. A generalizable model demonstrated robust performance in external validation. Prospective, multi-site assessment of a generalizable model coupled with brain-based biomarkers is warranted to assess the combined utility for identifying patients at high-risk for incurred neurologic morbidity and evaluating interventions to improve outcomes in this population.

## Supporting information

Supplemental Biomarker Methods

TRIPOD

## Data Availability

De-identified versions of data produced in the present study are available upon reasonable request to the authors.

## Acknowledgements

We would like to posthumously thank Dr. Ron Hayes (Banyan Biomarkers) for brain biomarker measurements; Dr. Pat Kochanek for helpful guidance; Dr. Henry Ogoe for helping to run an early version of the pipeline at UPMC Children’s Hospital of Pittsburgh; Mrs. Nassima Bouhenni for her initial efforts updating code; Mr. Dan Ricketts for his assistance setting up and maintaining virtual machines used for the final analyses; and Mr. Thomas Mathie for his assistance querying EHR data. United States Patent Application No. 17/760,558 and International Patent Application PCT/US2020/061985 have been filed related to this work.

## Data Sharing Statement

Data are available for specific use cases with investigator approval.

## Funding

NINDS R01NS118716 and NLM 5T15LM007059-38

## Additional Materials

### Supplemental Model Methods

**Supplemental Table 1.**
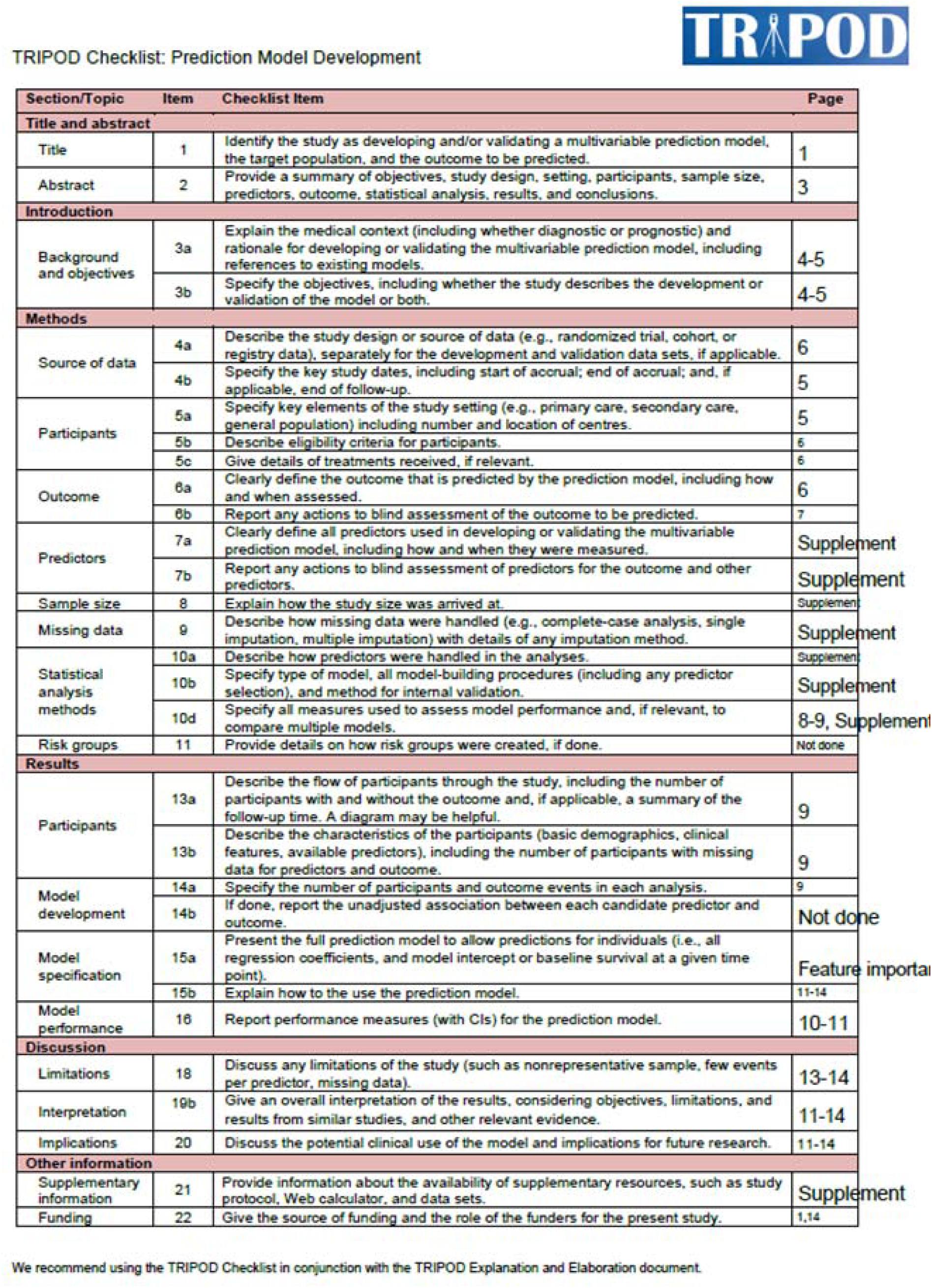
TRIPOD checklist.

**Supplemental Table 2.**
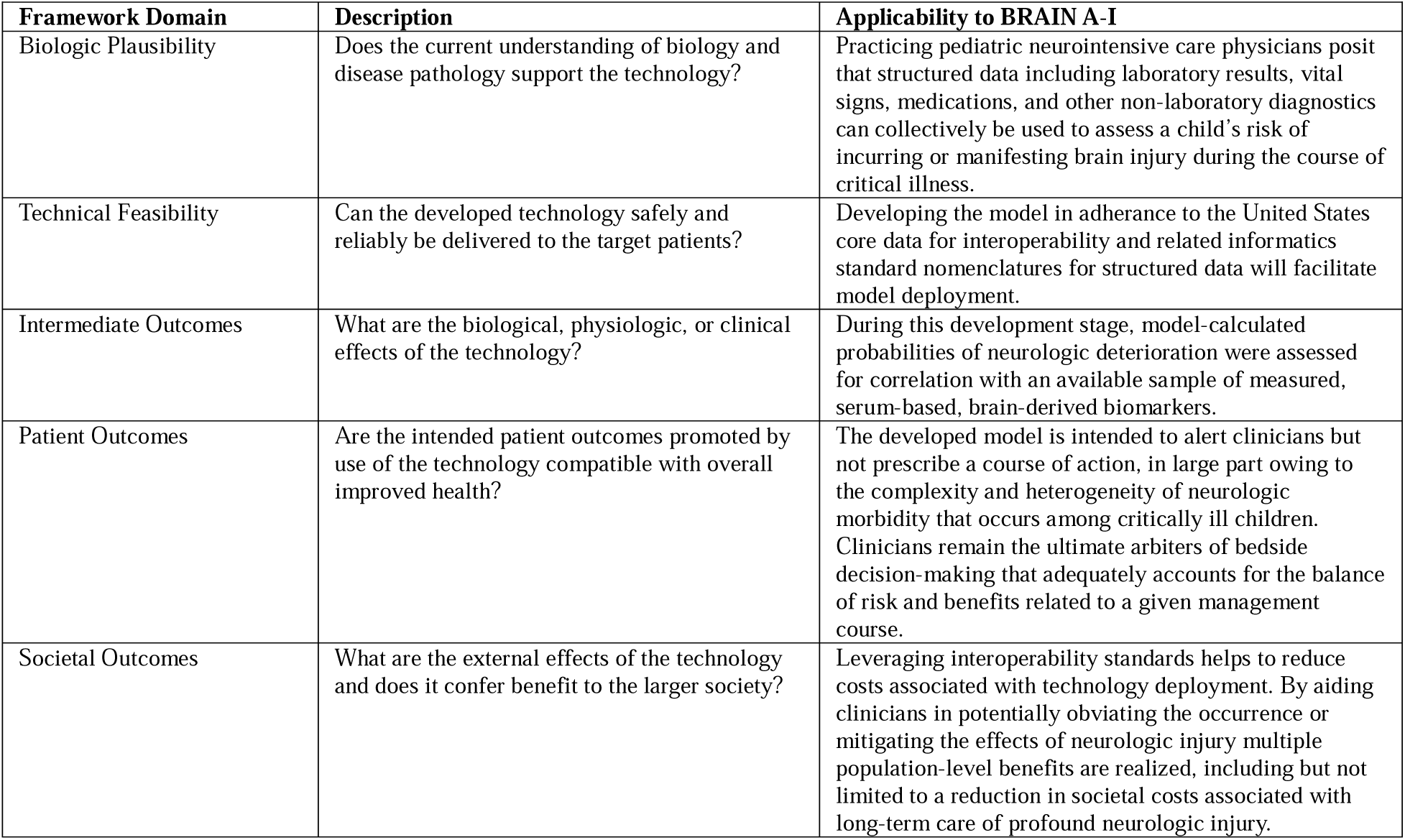
Littenberg framework for the assessment of medical technology as applied to BRAIN A-I.

**Supplemental Table 3.**
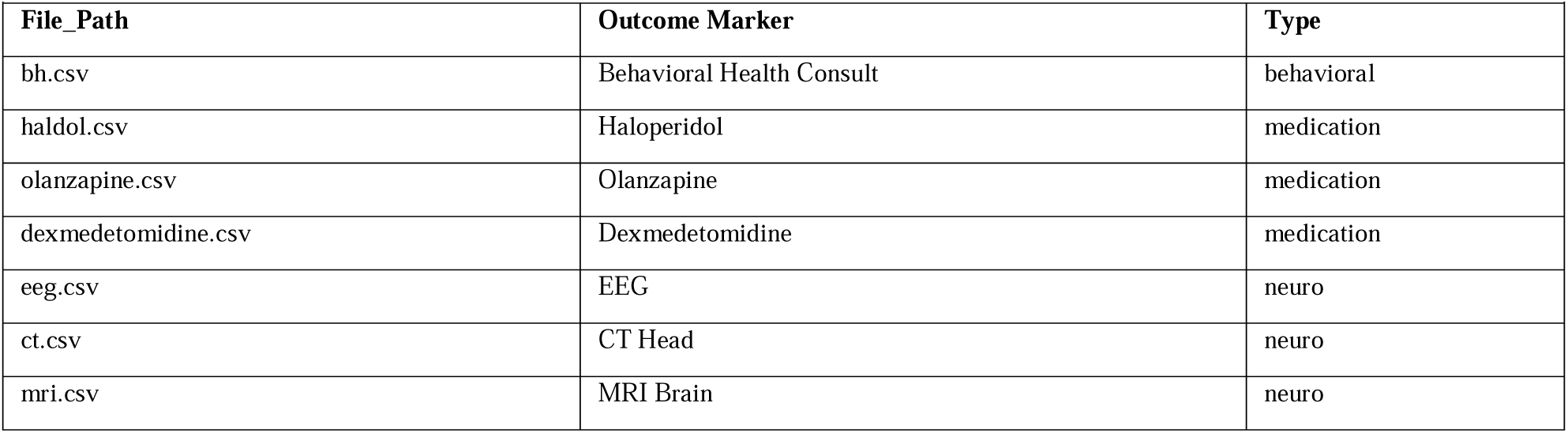
Data curation steps for individual data elements for the BRAIN AI outcome.

**Supplemental Table 4.** Data curation steps for individual data elements for BRAIN A-I.

| Data Element | Min | Max | Action | Type | Available at Both Sites |
| --- | --- | --- | --- | --- | --- |
| Base deficit | -30 | 0 | Discard | Numerical | Yes |
| Base excess | 0 | 30 | Discard | Numerical | Yes |
| Bicarbonate | 0 | 80 | Discard | Numerical | Yes |
| Blood urea nitrogen | 0 | 200 | Truncate | Numerical | Yes |
| Chloride | 60 | 190 | Discard | Numerical | Yes |
| Cisatracurium | 0 | 1 | Ignore | Boolean | Yes |
| C-Reactive Protein | 0 | 100 | Discard | Numerical | Yes |
| Creatinine | 0.1 | 25 | Discard | Numerical | Yes |
| CRRT Therapy Type | 0 | 1 | Discard | Boolean | No |
| DBP | 0 | 200 | Discard | Numerical | Yes |
| Dobutamine | 0 | 1 | Ignore | Boolean | Yes |
| Dopamine | 0 | 1 | Ignore | Boolean | Yes |
| ECMO Type | 0 | 1 | Discard | Boolean | No |
| Endotracheal tube | 0 | 1 | Discard | Boolean | No |
| Epinephrine | 0 | 1 | Ignore | Boolean | Yes |
| Fentanyl | 0 | 1 | Ignore | Boolean | Yes |
| Glucose | 0 | 2000 | Discard | Numerical | Yes |
| Hemoglobin | 0 | 30 | Discard | Numerical | Yes |
| Hydromorphone | 0 | 1 | Ignore | Boolean | Yes |
| INR | 0 | 25 | Discard | Numerical | No |
| Lactate | 0 | 30 | Discard | Numerical | Yes |
| Lorazepam | 0 | 1 | Ignore | Numerical | Yes |
| MBP | 0 | 160 | Discard | Numerical | Yes |
| Midazolam | 0 | 1 | Ignore | Boolean | Yes |
| Milrinone | 0 | 1 | Ignore | Boolean | Yes |
| Morphine | 0 | 1 | Ignore | Boolean | Yes |
| Norepinephrine | 0 | 1 | Ignore | Boolean | Yes |
| pCO2 | 5 | 150 | Discard | Numerical | Yes |
| Peds Coma Score | 3 | 15 | Discard | Numerical | Yes |
| pH | 6 | 8 | Discard | Numerical | Yes |
| Platelets | 0 | 5000 | Discard | Numerical | Yes |
| Potassium | 0.05 | 12 | Discard | Numerical | Yes |
| Procalcitonin | 0 | 250 | Discard | Numerical | Yes |
| PTT | 0 | 250 | Truncate | Numerical | Yes |
| Pulse | 0 | 350 | Discard | Numerical | Yes |
| Pupillary Reaction |  |  |  | Categorical | Yes |
| Respiratory Rate | 0 | 150 | Discard | Numerical | Yes |
| SBP | 0 | 300 | Discard | Numerical | Yes |
| Sodium | 80 | 215 | Discard | Numerical | Yes |
| SpO2 | 0 | 100 | Discard | Numerical | Yes |
| Temperature | 0 | 46 | Discard | Numerical | Yes |
| Ventilated | 0 | 1 | Discard | Boolean | Yes |
| Ventilator Make/Model |  |  |  | Categorical | Yes |
| Weight | 0 | 300 | Discard | Numerical | Yes |
| White blood cell count | 0 | 300 | Discard | Numerical | Yes |

**Supplemental Table 5.** Standard vocabulary crosswalk for BRAIN-AI components.

|  |  |  |  |  |  |
| --- | --- | --- | --- | --- | --- |
| BRAIN A-I Components | Cerner Millennium Code Set | Standard Vocabulary | Standard Identifier | Standard Display | USCDI |
| Laboratory Tests |  |  |  |  |  |
| Base deficit | 72 (Clinical Event Observation) | LOINC | 1922-4 | Base deficit in Arterial blood | Version 1 |
|  |  |  | 1923-2 | Base deficit in Capillary blood |  |
|  |  |  | 1924-0 | Base deficit in Venous blood |  |
|  |  |  | 30318-0 | Base deficit in Blood |  |
| Base excess | 72 (Clinical Event Observation) | LOINC | 11555-0 | Base excess in Blood by calculation | Version 1 |
|  |  |  | 1925-7 | Base excess in Arterial blood by calculation |  |
|  |  |  | 1926-5 | Base excess in Capillary blood by calculation |  |
|  |  |  | 1927-3 | Base excess in Venous blood by calculation |  |
| Bicarbonate | 72 (Clinical Event Observation) | LOINC | 1959-6 | Bicarbonate [Moles/volume] in Blood | Version 1 |
|  |  |  | 1960-4 | Bicarbonate [Moles/volume] in Arterial blood |  |
|  |  |  | 1961-2 | Bicarbonate [Moles/volume] in Capillary blood |  |
|  |  |  | 14627-4 | Bicarbonate [Moles/volume] in Venous blood |  |
|  |  |  | 2028-9 | Carbon dioxide, total [Moles/volume] in Serum or Plasma |  |
|  |  |  | 20565-8 | Carbon dioxide, total [Moles/volume] in Blood |  |
| Blood urea nitrogen | 72 (Clinical Event Observation) | LOINC | 3094-0 | Urea nitrogen [Mass/volume] in Serum or Plasma | Version 1 |
|  |  |  | 6299-2 | Urea nitrogen [Mass/volume] in Blood |  |
| Chloride | 72 (Clinical Event Observation) | LOINC | 2075-0 | Chloride [Moles/volume] in Serum or Plasma | Version 1 |
|  |  |  | 2069-3 | Chloride [Moles/volume] in Blood |  |
| C-reactive protein | 72 (Clinical Event Observation) | LOINC | 1988-5 | C reactive protein [Mass/volume] in Serum or Plasma | Version 1 |
| Creatinine | 72 (Clinical Event Observation) | LOINC | 2160-0 | Creatinine [Mass/volume] in Serum or Plasma | Version 1 |
|  |  |  | 38483-4 | Creatinine [Mass/volume] in Blood |  |
| Glucose | 72 (Clinical Event Observation) | LOINC | 41653-7 | Glucose [Mass/volume] in Capillary blood by Glucometer | Version 1 |
|  |  |  | 2345-7 | Glucose [Mass/volume] in Serum or Plasma |  |
|  |  |  | 2339-0 | Glucose [Mass/volume] in Blood |  |
| Hemoglobin | 72 (Clinical Event Observation) | LOINC | 718-7 | Hemoglobin [Mass/volume] in Blood | Version 1 |
|  |  |  | 30351-1 | Hemoglobin [Mass/volume] in Mixed venous blood |  |
|  |  |  | 30313-1 | Hemoglobin [Mass/volume] in Arterial blood |  |
|  |  |  | 30350-3 | Hemoglobin [Mass/volume] in Venous blood |  |
| International normalized ratio | 72 (Clinical Event Observation) | LOINC | 6301-6 | INR in Platelet poor plasma by Coagulation assay | Version 1 |
| Lactate | 72 (Clinical Event Observation) | LOINC | 2519-7 | Lactate [Moles/volume] in Venous blood | Version 1 |
|  |  |  | 32693-4 | Lactate [Moles/volume] in Blood |  |
| Partial pressure of carbon dioxide | 72 (Clinical Event Observation) | LOINC | 2020-6 | Carbon dioxide [Partial pressure] in Capillary blood | Version 1 |
|  |  |  | 11557-6 | Carbon dioxide [Partial pressure] in Blood |  |
|  |  |  | 2019-8 | Carbon dioxide [Partial pressure] in Arterial blood |  |
| pH | 72 (Clinical Event Observation) | LOINC | 2745-8 | pH of Capillary blood | Version 1 |
|  |  |  | 11558-4 | pH of Blood |  |
|  |  |  | 2744-1 | pH of Arterial blood |  |
|  |  |  | 2746-6 | pH of Venous blood |  |
| Platelets | 72 (Clinical Event Observation) | LOINC | 777-3 | Platelets [# /volume] in Blood by Automated count | Version 1 |
| BRAIN A-I Components | Cerner Millennium Code Set | Standard Vocabulary | Standard Identifier | Standard Display | USCDI |
| Potassium | 72 (Clinical Event Observation) | LOINC | 2823-3 | Potassium [Moles/volume] in Serum or Plasma | Version 1 |
|  |  |  | 6298-4 | Potassium [Moles/volume] in Blood |  |
| Procalcitonin | 72 (Clinical Event Observation) | LOINC | 33959-8 | Procalcitonin [Mass/volume] in Serum or Plasma | Version 1 |
| Partial thromboplastin time | 72 (Clinical Event Observation) | LOINC | 14979-9 | aPTT in Platelet poor plasma by Coagulation assay | Version 1 |
| Sodium | 72 (Clinical Event Observation) | LOINC | 2951-2 | Sodium [Moles/volume] in Serum or Plasma | Version 1 |
|  |  |  | 2947-0 | Sodium [Moles/volume] in Blood |  |
| White blood cell count | 72 (Clinical Event Observation) | LOINC | 6690-2 | Leukocytes [# /volume] in Blood by Automated count | Version 1 |
|  |  |  | 49498-9 | Leukocytes [# /volume] in Blood by Estimate |  |
| Vital signs |  |  |  |  |  |
| Diastolic blood pressure | 72 (Clinical Event Observation) | LOINC | 8453-3 | Diastolic blood pressure--sitting | Version 1 |
|  |  |  | 8454-1 | Diastolic blood pressure--standing |  |
|  |  |  | 8455-8 | Diastolic blood pressure--supine |  |
|  |  |  | 8462-4 | Diastolic blood pressure |  |
| Glasgow coma scale score | 72 (Clinical Event Observation) | LOINC | 9269-2 | Glasgow coma score total | Version 3 |
|  |  |  | 9270-0 | Glasgow coma score verbal |  |
|  |  |  | 9267-6 | Glasgow coma score eye opening |  |
|  |  |  | 9268-4 | Glasgow coma score motor |  |
| Heart rate | 72 (Clinical Event Observation) | LOINC | 68999-2 | Heart rate --supine | Version 1 |
|  |  |  | 69000-8 | Heart rate --sitting |  |
|  |  |  | 69001-6 | Heart rate --standing |  |
|  |  |  | 8867-4 | Heart rate |  |
|  |  |  | 8890-6 | Heart rate Cardiac apex by Auscultation |  |
| Mean blood pressure | 72 (Clinical Event Observation) | LOINC | 8478-0 | Mean blood pressure | Version 1 |
| Pulse oximetry | 72 (Clinical Event Observation) | LOINC | 59408-5 | Oxygen saturation in Arterial blood by Pulse oximetry | Version 1 |
| Respiratory rate | 72 (Clinical Event Observation) | LOINC | 9279-1 | Respiratory rate | Version 1 |
| Systolic blood pressure | 72 (Clinical Event Observation) | LOINC | 8480-6 | Systolic blood pressure | Version 1 |
| Temperature | 72 (Clinical Event Observation) | LOINC | 60836-4 | Esophageal temperature | Version 1 |
|  |  |  | 76278-1 | Bladder temperature via Foley |  |
|  |  |  | 8310-5 | Body temperature |  |
|  |  |  | 8328-7 | Axillary temperature |  |
|  |  |  | 8329-5 | Body temperature - Core |  |
|  |  |  | 8331-1 | Oral temperature |  |
|  |  |  | 8332-9 | Rectal temperature |  |
|  |  |  | 8334-5 | Body temperature – Urinary bladder |  |
| Ventilator interface | 72 (Clinical Event Observation) | LOINC | LL5542-7 | Intubation tube types | Medical Device Class - Version Unknown |
| Ventilator make | 72 (Clinical Event Observation) | LOINC | LL7706-7 | Ventilator | Medical Device Class - Version Unknown |
| Weight | 72 (Clinical Event Observation) | LOINC | 29463-7 | Body weight | Version 1 |
| Medications |  |  |  |  |  |
| Cisatracurium (Intravenous) | 72 (Clinical Event Observation) | RxNorm RXCUI | 319864 | cisatracurium | Version 1 |
| Dexmedetomidine (Intravenous) | 72 (Clinical Event Observation) | RxNorm RXCUI | 48937 | dexmedeTOMIDine | Version 1 |
| BRAIN A-I Components | Cerner Millennium Code Set | Standard Vocabulary | Standard Identifier | Standard Display | USCDI |
| Dobutamine (Intravenous) | 72 (Clinical Event Observation) | RxNorm RXCUI | 3616 | DOBU**T**amine | Version 1 |
| Dopamine (Intravenous) | 72 (Clinical Event Observation) | RxNorm RXCUI | 3628 | dopa**m**ine | Version 1 |
| Epinephrine (Intravenous) | 72 (Clinical Event Observation) | RxNorm RXCUI | 3992 | EPINEPH**r**ine | Version 1 |
| Fentanyl (Intravenous) | 72 (Clinical Event Observation) | RxNorm RXCUI | 4337 | fenta**N**YL | Version 1 |
| Haldol | 72 (Clinical Event Observation) | RxNorm RXCUI | 151839 | Haldol | Version 1 |
| Hydromorphone (Intravenous) | 72 (Clinical Event Observation) | RxNorm RXCUI | 3423 | HYDRO**m**orphone | Version 1 |
| Lorazepam (Intravenous) | 72 (Clinical Event Observation) | RxNorm RXCUI | 6470 | LORazepa**m** | Version 1 |
| Midazolam (Intravenous) | 72 (Clinical Event Observation) | RxNorm RXCUI | 6960 | midazolam | Version 1 |
| Milrinone (Intravenous) | 72 (Clinical Event Observation) | RxNorm RXCUI | 52769 | milrinone | Version 1 |
| Morphine (Intravenous) | 72 (Clinical Event Observation) | RxNorm RXCUI | 7052 | morphine | Version 1 |
| Norepinephrine (Intravenous) | 72 (Clinical Event Observation) | RxNorm RXCUI | 7512 | norepinephrine | Version 1 |
| Olanzapine | 72 (Clinical Event Observation) | RxNorm RXCUI | 61381 | OLANZapine | Version 1 |
| Non-Laboratory Diagnostics |  |  |  |  |  |
| Brain computed tomography | 72 (Clinical Event Observation) | LOINC | 24725-4 | CT Head | Version 2 |
| Brain magnetic resonance imaging | 72 (Clinical Event Observation) | LOINC | 24590-2 | MR Brain | Version 2 |
| Electroencephalogram | 72 (Clinical Event Observation) | LOINC | 11523-8 | EEG study | Version 2 |
| Pupillary reaction | 72 (Clinical Event Observation) | LOINC | 79899-1 | Left pupil Pupillary response | Observation Class – Version Unknown |
|  |  | LOINC | 79815-7 | Right pupil Pupillary response | Observation Class – Version Unknown |
| Consultation |  |  |  |  |  |
| Behavioral health consult | 72 (Clinical Event Observation) | SNOMED CT | 733870009 | Assessment of delirium | Observation Class – Version Unknown |
| Abbreviations: aPTT, activated partial thromboplastin time; BRAIN A-I, biodigital rapid alert for identifying neuromorbidity A-I bundle; CT, computed tomography; EEG, electroencephalography; MR, magnetic resonance; LOINC, logical object identifiers, names, and codes; RXCUI, RxNorm concept unique identifier; SNOMED CT, systematized nomenclature of medicine - clinical terms; USCDI, United States Core Data for Interoperability |  |  |  |  |  |

**Supplemental Table 6.** Predictive performance of the XGBoost model with a 12-hour censor horizon and 48-hour feature window in the test dataset, after manual tuning, and after Bayesian tuning.

| **Supplemental Table 6.** Predictive performance of the XGBoost model with a 12-hour censor horizon and 48-hour feature window in the validation dataset, after manual tuning, and after Bayesian tuning. |  |  |  |
| --- | --- | --- | --- |
|  | **XGBoost Base** | **XGBoost Manual Tuning** | **XGBoost Bayesian Tuning** |
| **AUROC** | 0.84 | 0.84 | 0.85 |
| **AUPRC** | 0.61 | 0.62 | 0.63 |
| **F1 Score** | 0.54 | 0.56 | 0.58 |
| **PPV** | 0.77 | 0.75 | 0.63 |
| **NPV** | 0.92 | 0.92 | 0.93 |
| **Sensitivity** | 0.41 | 0.44 | 0.54 |
| **Specificity** | 0.98 | 0.98 | 0.95 |
| Models were tuned to optimize the F1 score, bordered in bold.<br>Abbreviations: AUPRC, area under the precision recall curve; AUROC, area under the receiver operating characteristics curve; NPV, negative predictive value; PPV, positive predictive value; XGBoost, extreme gradient boosting. |  |  |  |

**Supplemental Table 7.** Predictive performance of the XGBoost model with a 12-hour censor horizon and 48-hour feature window in the validation dataset, after manual tuning, and after Bayesian tuning.

| **Supplemental Table 7.** Predictive performance of the XGBoost model with a 12-hour censor horizon and 48-hour feature window in the test dataset, after manual tuning, and after Bayesian tuning. |  |  |  |
| --- | --- | --- | --- |
|  | **XGBoost Base** | **XGBoost Manual Tuning** | **XGBoost Bayesian Tuning** |
| **AUROC** | 0.87 | 0.88 | 0.89 |
| **AUPRC** | 0.66 | 0.68 | 0.69 |
| **F1 Score** | 0.57 | 0.58 | 0.62 |
| **PPV** | 0.86 | 0.79 | 0.74 |
| **NPV** | 0.93 | 0.93 | 0.94 |
| **Sensitivity** | 0.43 | 0.46 | 0.53 |
| **Specificity** | 0.99 | 0.98 | 0.98 |
| Models were tuned to optimize the F1 score, bordered in bold.<br>Abbreviations: AUPRC, area under the precision recall curve; AUROC, area under the receiver operating characteristics curve; NPV, negative predictive value; PPV, positive predictive value; XGBoost, extreme gradient boosting. |  |  |  |

**Supplemental Table 8.** Cohort ascertainment and exclusions for varied censored time horizons and feature windows at the development site.

|  |  |  |  |  |  |  |  |  |  |
| --- | --- | --- | --- | --- | --- | --- | --- | --- | --- |
| Encounters with a PICU admission | **32,702** |  |  |  |  |  |  |  |  |
| Feature window (hours) | 24 |  |  | 48 |  |  | 72 |  |  |
| Censored time Horizon (hours) | 6 | 12 | 24 | 6 | 12 | 24 | 6 | 12 | 24 |
| Visit length or time to outcome hours less than feature window + time horizon | 2925 | 3435 | 3908 | 7720 | 8230 | 8703 | 11593 | 12103 | 12576 |
| Outcome occurred before PICU admission | 4956 |  |  |  |  |  |  |  |  |
| Missing age, admission time, or discharge time | 821 |  |  |  |  |  |  |  |  |
| PICU length of stay <1 hour | 182 |  |  |  |  |  |  |  |  |
| No documented SpO<sub>2</sub> measurement | 7 |  |  |  |  |  |  |  |  |
| Discharge time documented prior to admission time | 2 |  |  |  |  |  |  |  |  |
| Final cohort | **23873** | **23363** | **22890** | **19078** | **18568** | **18095** | **15205** | **14695** | **14222** |
| Cases | **2841** | **2331** | **1858** | **2841** | **2331** | **1858** | **2841** | **2331** | **1858** |
| Controls | **21032** |  |  | **16237** |  |  | **12364** |  |  |

Cohort numbers in **bold** represent the cohort at a given stage of ascertainment and numbers not in bold represent encounter dropout at stages of cleaning.

**Supplemental Table 9.** Performance of the optimal models in the validation dataset at the development and validation sites.

|  | Development Site |  |  |  |  |  |  |  |  |
| --- | --- | --- | --- | --- | --- | --- | --- | --- | --- |
| Feature Window (hours) | 24 |  |  | 48 |  |  | 72 |  |  |
| Censored Time Horizon (hours) | 6 | 12 | 24 | 6 | 12 | 24 | 6 | 12 | 24 |
| Model | LR | LR | LR | XGB | XGB | LR | XGB | LR | XGB |
| Feature Selection | IG | IG | IG | IG | IG | IG | IG | IG | IG |
| AUROC | 0.83 | 0.83 | 0.80 | 0.87 | 0.82 | 0.82 | 0.88 | 0.86 | 0.82 |
| AUPRC | 0.59 | 0.49 | 0.39 | 0.73 | 0.61 | 0.53 | 0.78 | 0.71 | 0.61 |
| PPV | 0.74 | 0.65 | 0.60 | 0.82 | 0.79 | 0.69 | 0.88 | 0.75 | 0.68 |
| NPV | 0.92 | 0.92 | 0.93 | 0.92 | 0.92 | 0.93 | 0.91 | 0.92 | 0.92 |
| Sensitivity | 0.40 | 0.29 | 0.24 | 0.53 | 0.41 | 0.41 | 0.58 | 0.54 | 0.47 |
| Specificity | 0.98 | 0.98 | 0.99 | 0.98 | 0.98 | 0.98 | 0.98 | 0.97 | 0.97 |
| Abbreviations: AUROC, area under the receiver operating characteristics curve; AUPRC, area under the precision recall curve; CFS, correlation-based feature selection; IG, information gain; LR, logistic regression; NB, naïve Bayes; NPV, negative predictive value; PPV, positive predictive value; XGB, XGBoost (extreme gradient boosting) |  |  |  |  |  |  |  |  |  |

**Supplemental Table 10.** **A)** F1 scores of top performing models in the development site validation dataset. **B)** F1 scores of top performing models in the validation site test dataset.

|  |  | Censored Time Horizon |  |  |
| --- | --- | --- | --- | --- |
|  |  | 6-hours | 12-hours | 24-hours |
| Feature Window | 24-hours | LR<br>0.52 | LR<br>0.40 | LR<br>0.34 |
|  | 48-hours | XGB<br>0.65 | XGB<br>0.54 | LR<br>0.51 |
|  | 72-hours | XGB<br>0.70 | LR<br>0.65 | XGB<br>0.57 |
| Cells denote the best performing model and the F1 score / Brier score.<br>All models at the development site were created using information gain feature selection<br>Abbreviations: LR, logistic regression; XGB, XGBoost (extreme gradient boosting) |  |  |  |  |

**Supplemental Table 11.**
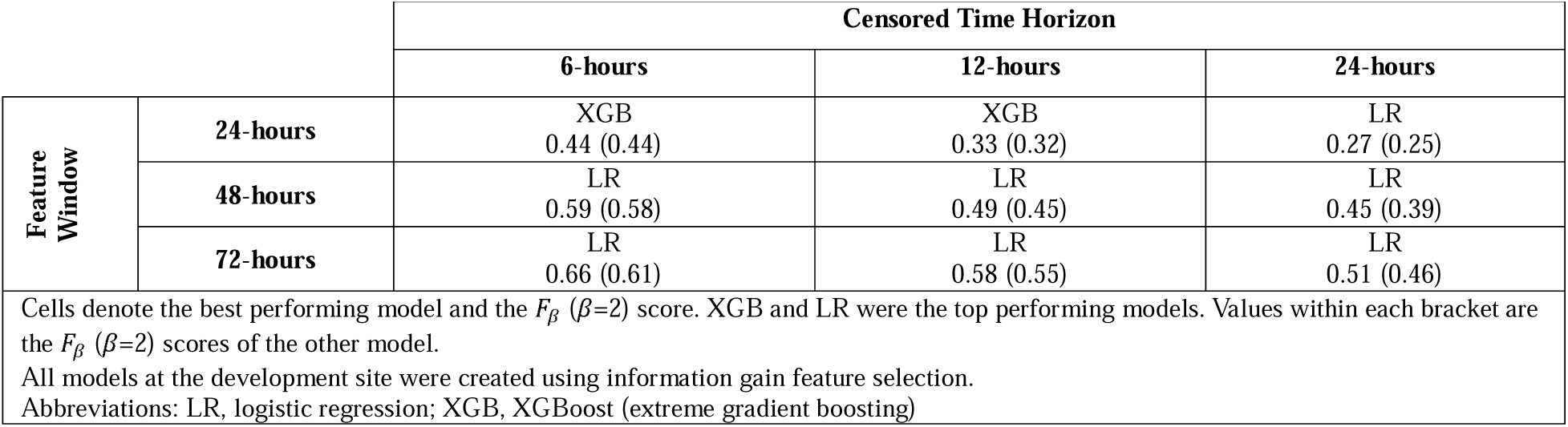
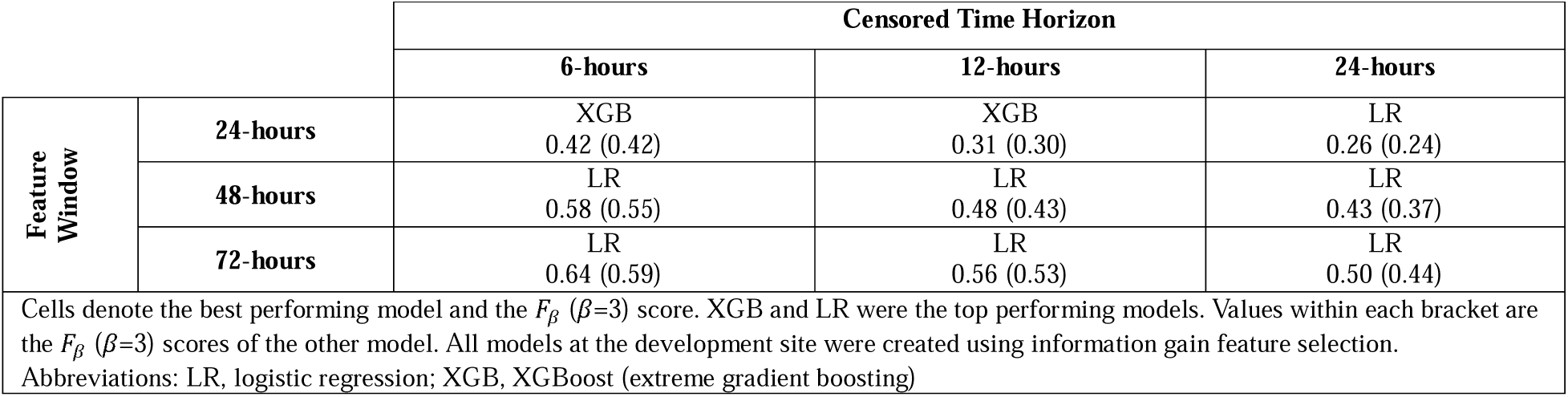
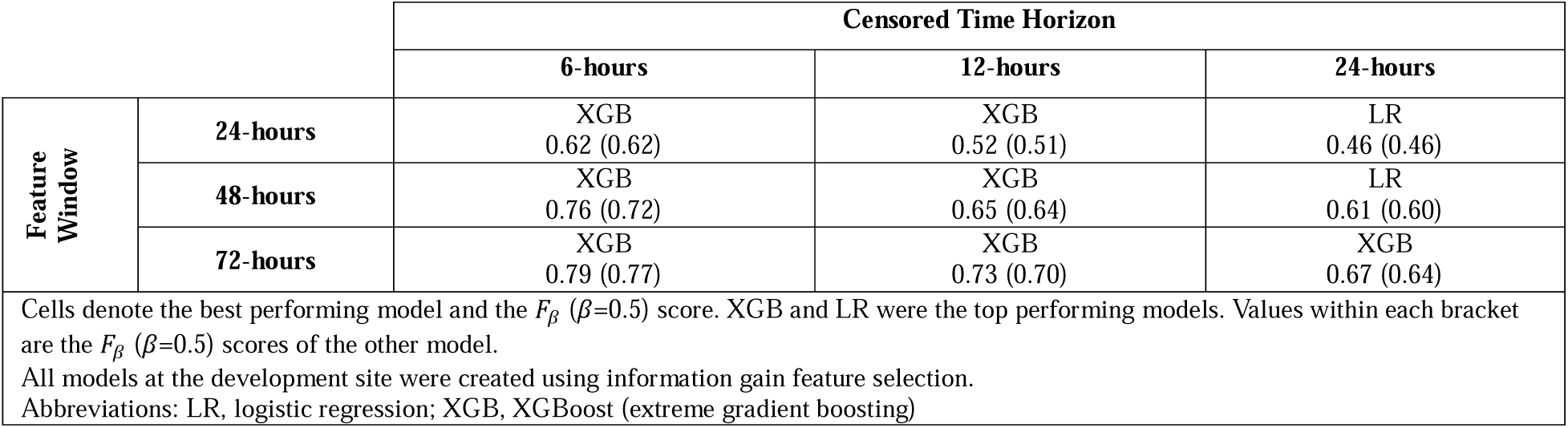
**A)** F_β_ (β = 2) scores of top performing models in the development site validation dataset; **B**) F_β_ (β = 3) scores of top performing models in the development site validation dataset; **C**) F_β_ (β = 0.5) scores of top performing models in the development site validation dataset.

**Supplemental Table 12.** **A)** Statistical performance of the 12-hour time horizon, 48-hour feature window XGBoost and logistic regression models over a range of score thresholds in the development site validation dataset; **B**) Statistical performance of the 12-hour time horizon, 48-hour feature window XGBoost model over a range of score thresholds in the development site test dataset.

| Supplemental Table 12A. Statistical performance of the 12-hour time horizon, 48-hour feature window XGBoost and logistic regression models over a range of score thresholds in the development site validation dataset. |  |  |  |  |  |  |  |  |  |  |  |
| --- | --- | --- | --- | --- | --- | --- | --- | --- | --- | --- | --- |
| Threshold | AUROC | AUPRC | F1 Score | F2 Score | F3 Score | F0.5 Score | PPV | NPV | Sensitivity | Specificity | Confusion Matrix - (TN, FP, FN,TP) |
| XGBoost Model |  |  |  |  |  |  |  |  |  |  |  |
| 0.025 | 0.818 | 0.601 | 0.361 | 0.542 | 0.650 | 0.271 | 0.232 | 0.956 | 0.813 | 0.602 | (1879, 1242, 86, 375) |
| 0.05 |  |  | 0.422 | 0.554 | 0.619 | 0.341 | 0.302 | 0.945 | 0.701 | 0.761 | (2375, 746, 138, 323) |
| 0.1 |  |  | 0.483 | 0.554 | 0.582 | 0.429 | 0.399 | 0.938 | 0.614 | 0.863 | (2694, 427, 178, 283) |
| 0.3 |  |  | 0.553 | 0.508 | 0.494 | 0.607 | 0.649 | 0.926 | 0.482 | 0.962 | (3001, 120, 239, 222) |
| 0.5 |  |  | 0.536 | 0.454 | 0.432 | 0.654 | 0.766 | 0.919 | 0.412 | 0.981 | (3063, 58, 271, 190) |
| 0.7 |  |  | 0.508 | 0.411 | 0.386 | 0.664 | 0.836 | 0.913 | 0.364 | 0.989 | (3088, 33, 293, 168) |
| 0.9 |  |  | 0.429 | 0.325 | 0.301 | 0.629 | 0.915 | 0.904 | 0.280 | 0.996 | (3109, 12, 332, 129) |
| Logistic Regression Model |  |  |  |  |  |  |  |  |  |  |  |
| 0.025 | 0.827 | 0.610 | 0.318 | 0.518 | 0.656 | 0.229 | 0.193 | 0.966 | 0.894 | 0.449 | (1402, 1719, 49, 412) |
| 0.05 |  |  | 0.384 | 0.561 | 0.662 | 0.292 | 0.252 | 0.958 | 0.809 | 0.645 | (2012, 1109, 88, 373) |
| 0.1 |  |  | 0.452 | 0.574 | 0.631 | 0.372 | 0.333 | 0.947 | 0.701 | 0.793 | (2475, 646, 138, 323) |
| 0.3 |  |  | 0.549 | 0.538 | 0.535 | 0.561 | 0.568 | 0.931 | 0.531 | 0.940 | (2935, 186, 216, 245) |
| 0.5 |  |  | 0.557 | 0.494 | 0.477 | 0.638 | 0.707 | 0.924 | 0.460 | 0.972 | (3033, 88, 249, 212) |
| 0.7 |  |  | 0.533 | 0.444 | 0.420 | 0.668 | 0.803 | 0.917 | 0.399 | 0.986 | (3076, 45, 277, 184) |
| 0.9 |  |  | 0.450 | 0.346 | 0.321 | 0.645 | 0.908 | 0.906 | 0.299 | 0.996 | (3107, 14, 323, 138) |

| **Supplemental Table 12B.** Statistical performance of the 12-hour time horizon, 48-hour feature window XGBoost model over a range of score thresholds in the development site test dataset. |  |  |  |  |  |  |  |  |  |  |  |
| --- | --- | --- | --- | --- | --- | --- | --- | --- | --- | --- | --- |
| Threshold | AUROC | AUPRC | F1 Score | F2 Score | F3 Score | F0.5 Score | PPV | NPV | Sensitivity | Specificity | Confusion Matrix - (TN, FP, FN,TP) |
| **XGBoost Model** |  |  |  |  |  |  |  |  |  |  |  |
| 0.025 | 0.873 | 0.671 | 0.399 | 0.587 | **0.698** | 0.302 | 0.260 | **0.974** | **0.859** | 0.682 | (2559, 1195, 69, 419) |
| 0.05 |  |  | 0.493 | 0.621 | 0.679 | 0.409 | 0.367 | 0.962 | 0.750 | 0.832 | (3124, 630, 122, 366) |
| 0.1 |  |  | 0.573 | **0.623** | 0.642 | 0.531 | 0.505 | 0.954 | 0.662 | 0.916 | (3438, 316, 165, 323) |
| 0.3 |  |  | **0.603** | 0.538 | 0.519 | 0.685 | 0.754 | 0.938 | 0.502 | 0.979 | (3674, 80, 243, 245) |
| 0.5 |  |  | 0.562 | 0.463 | 0.437 | **0.715** | 0.874 | 0.929 | 0.414 | 0.992 | (3725, 29, 286, 202) |
| 0.7 |  |  | 0.491 | 0.383 | 0.357 | 0.684 | 0.926 | 0.920 | 0.334 | 0.997 | (3741, 13, 325, 163) |
| 0.9 |  |  | 0.390 | 0.287 | 0.264 | 0.610 | **0.975** | 0.910 | 0.244 | **0.999** | (3751, 3, 369, 119) |

**Supplemental Table 13.** Cohort ascertainment and exclusions for varied feature windows for the validation site.

|  |  |
| --- | --- |
| Timeframe | **4/2018-2023** |
| Encounters with a PICU admission | **9,039** |
| Feature Window | 48-hours |
| Visit length <24 hours or PICU length of stay <1 hour | 1,791 |
| Missing age, discharge time or sex | 329 |
| No documented SpO<sub>2</sub> measurement | 15 |
| No accurate Neuro Consultation records prior 2018 | --- |
| Outcome event prior to PICU admission | 79 |
| Key data missing within Feature Window | --- |
| Final Cohort | **6,825** |
| Cases | **387** |
| Controls | **6,438** |

**Supplemental Table 14.** **A)** Statistical performance of the extreme gradient boosting (XGBoost) and logistic regression generalizable models at the development site across varied output thresholds. **B)** Statistical performance of the extreme gradient boosting (XGBoost) and logistic regression generalizable models at the external validation site across varied output thresholds.

| Threshold | AUROC | AUPRC | F1 Score | F2 Score | F3 Score | F0.5 Score | PPV | NPV | Sensitivity | Specificity | Confusion Matrix - (TN, FP, FN,TP) |
| --- | --- | --- | --- | --- | --- | --- | --- | --- | --- | --- | --- |
| XGBoost Model |  |  |  |  |  |  |  |  |  |  |  |
| 0.025 | 0.872 | 0.617 | 0.365 | 0.551 | 0.665 | 0.272 | 0.233 | 0.976 | 0.837 | 0.708 | (2658, 1096, 65, 333) |
| 0.05 |  |  | 0.459 | 0.598 | 0.666 | 0.372 | 0.330 | 0.970 | 0.751 | 0.838 | (3147, 607, 99, 299) |
| 0.1 |  |  | 0.528 | 0.591 | 0.615 | 0.478 | 0.450 | 0.960 | 0.641 | 0.917 | (3442, 312, 143, 255) |
| 0.3 |  |  | 0.547 | 0.481 | 0.462 | 0.635 | 0.711 | 0.943 | 0.445 | 0.981 | (3682, 72, 221, 177) |
| 0.5 |  |  | 0.489 | 0.394 | 0.370 | 0.645 | 0.818 | 0.935 | 0.349 | 0.992 | (3723, 31, 259, 139) |
| 0.7 |  |  | 0.421 | 0.316 | 0.292 | 0.629 | 0.939 | 0.928 | 0.271 | 0.998 | (3747, 7, 290, 108) |
| 0.9 |  |  | 0.312 | 0.222 | 0.202 | 0.527 | 0.974 | 0.921 | 0.186 | 0.999 | (3752, 2, 324, 74) |
| Logistic Regression Model |  |  |  |  |  |  |  |  |  |  |  |
| 0.025 | 0.855 | 0.605 | 0.303 | 0.497 | 0.633 | 0.218 | 0.183 | 0.977 | 0.869 | 0.590 | (2213, 1541, 52, 346) |
| 0.05 |  |  | 0.388 | 0.552 | 0.643 | 0.299 | 0.260 | 0.969 | 0.769 | 0.767 | (2881, 873, 92, 306) |
| 0.1 |  |  | 0.481 | 0.571 | 0.609 | 0.415 | 0.380 | 0.960 | 0.653 | 0.887 | (3330, 424, 138, 260) |
| 0.3 |  |  | 0.562 | 0.513 | 0.499 | 0.621 | 0.668 | 0.947 | 0.485 | 0.974 | (3658, 96, 205, 193) |
| 0.5 |  |  | 0.520 | 0.429 | 0.406 | 0.658 | 0.801 | 0.938 | 0.384 | 0.990 | (3716, 38, 245, 153) |
| 0.7 |  |  | 0.489 | 0.383 | 0.357 | 0.677 | 0.911 | 0.934 | 0.334 | 0.997 | (3741, 13, 265, 133) |
| 0.9 |  |  | 0.367 | 0.267 | 0.245 | 0.584 | 0.968 | 0.924 | 0.226 | 0.999 | (3751, 3, 308, 90) |

**Supplemental Figure 1.**
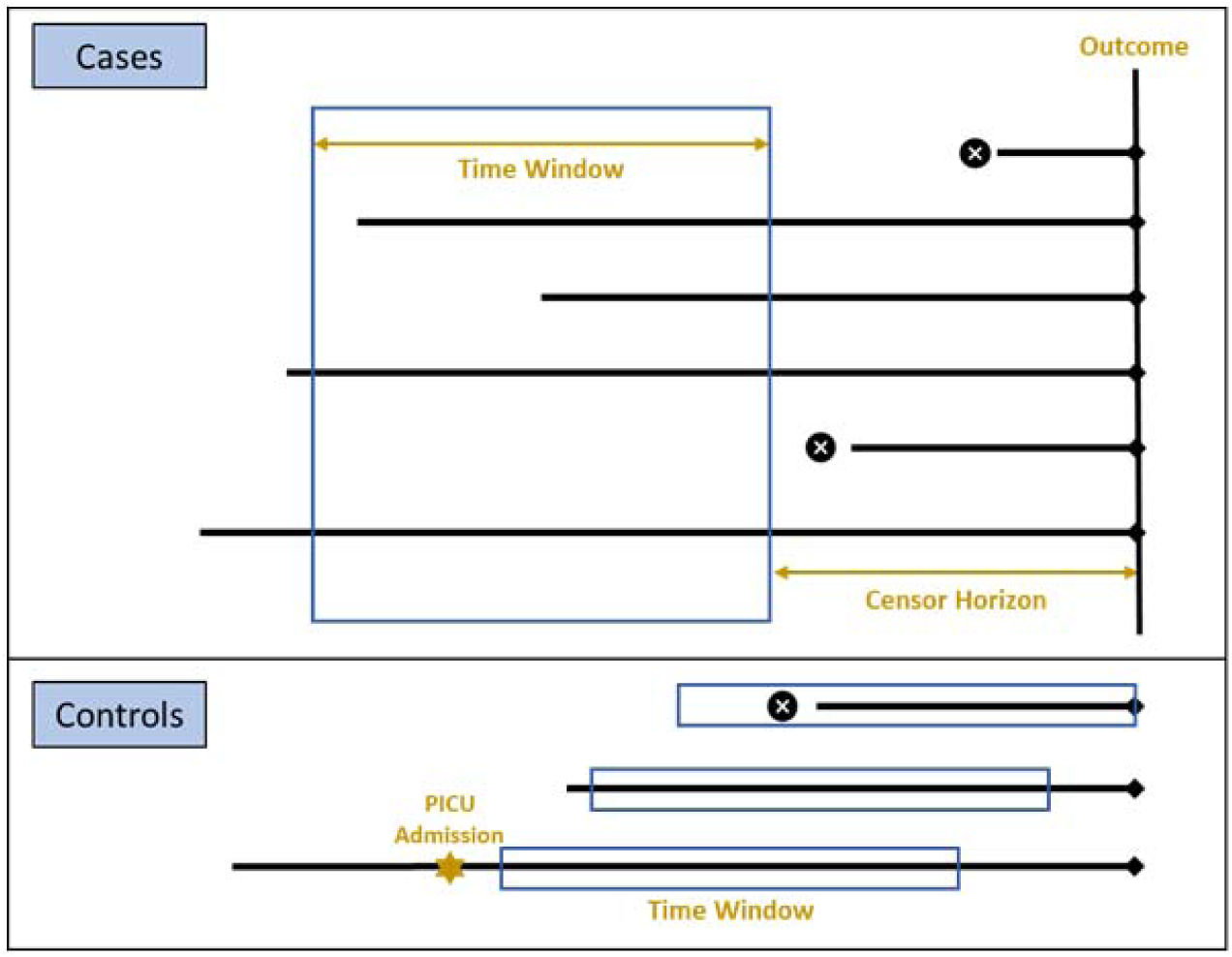
A representation of the time window and censor horizons used to define cases and controls as part of the development, validation, and test cohorts. The blue boxes in the top, ‘Cases’ box identify a time window that is also demarcated by a horizontal, gold, bidirectional arrow, the horizontal black lines represent length of stay for individual encounters, the vertical black line represents the occurrence of the neurological morbidity outcome, and the gold, horizontal, bidirectional arrow indicates the censor horizon, or period of time that data were not incorporated into the model. In the bottom, ‘Controls’ box, the blue boxes indicate the time window of data used for each stage of model development and evaluation.

**Supplemental Figure 2.**
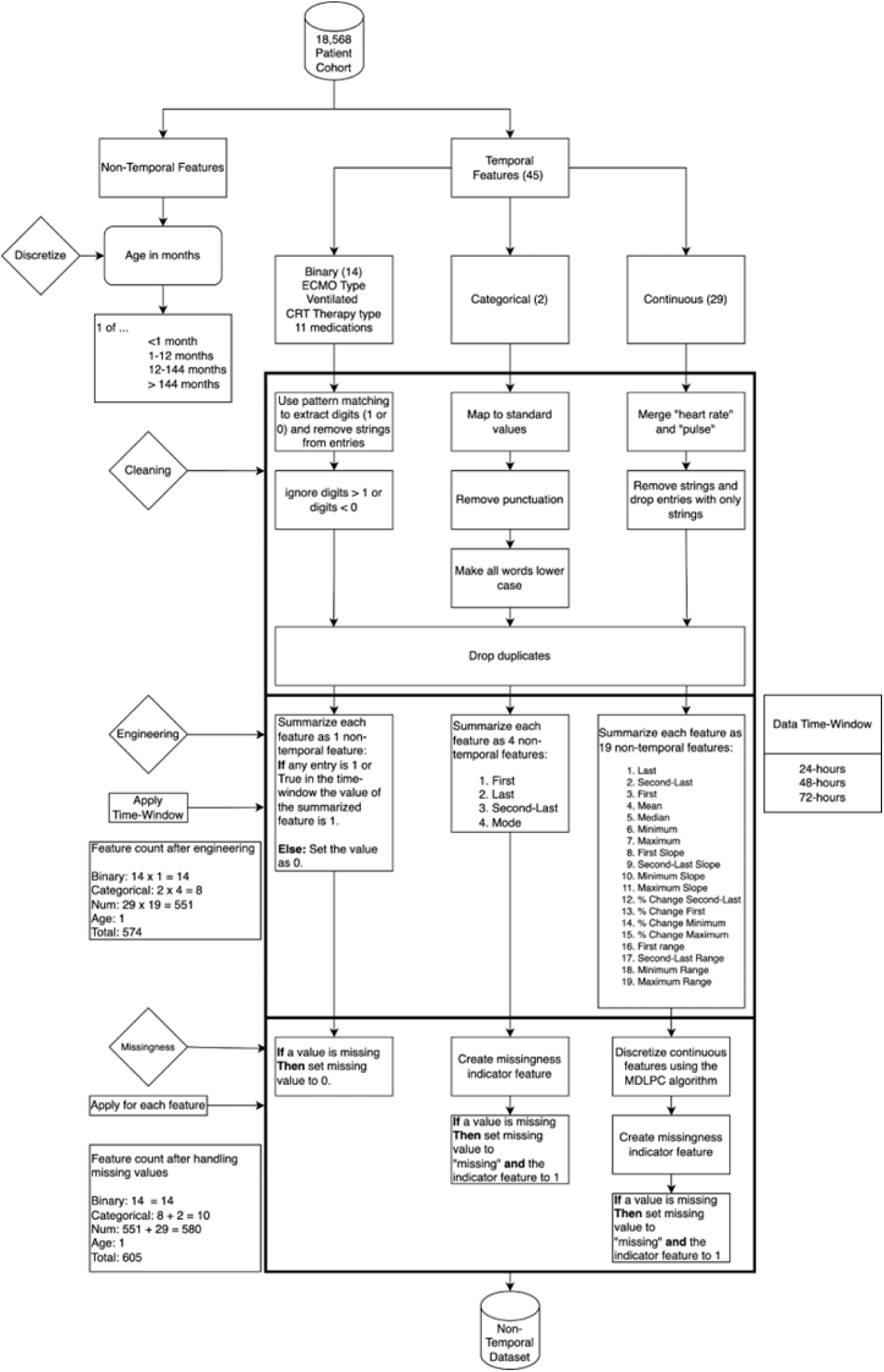
Data cleaning and feature engineering process.

**Supplemental Figure 3.**
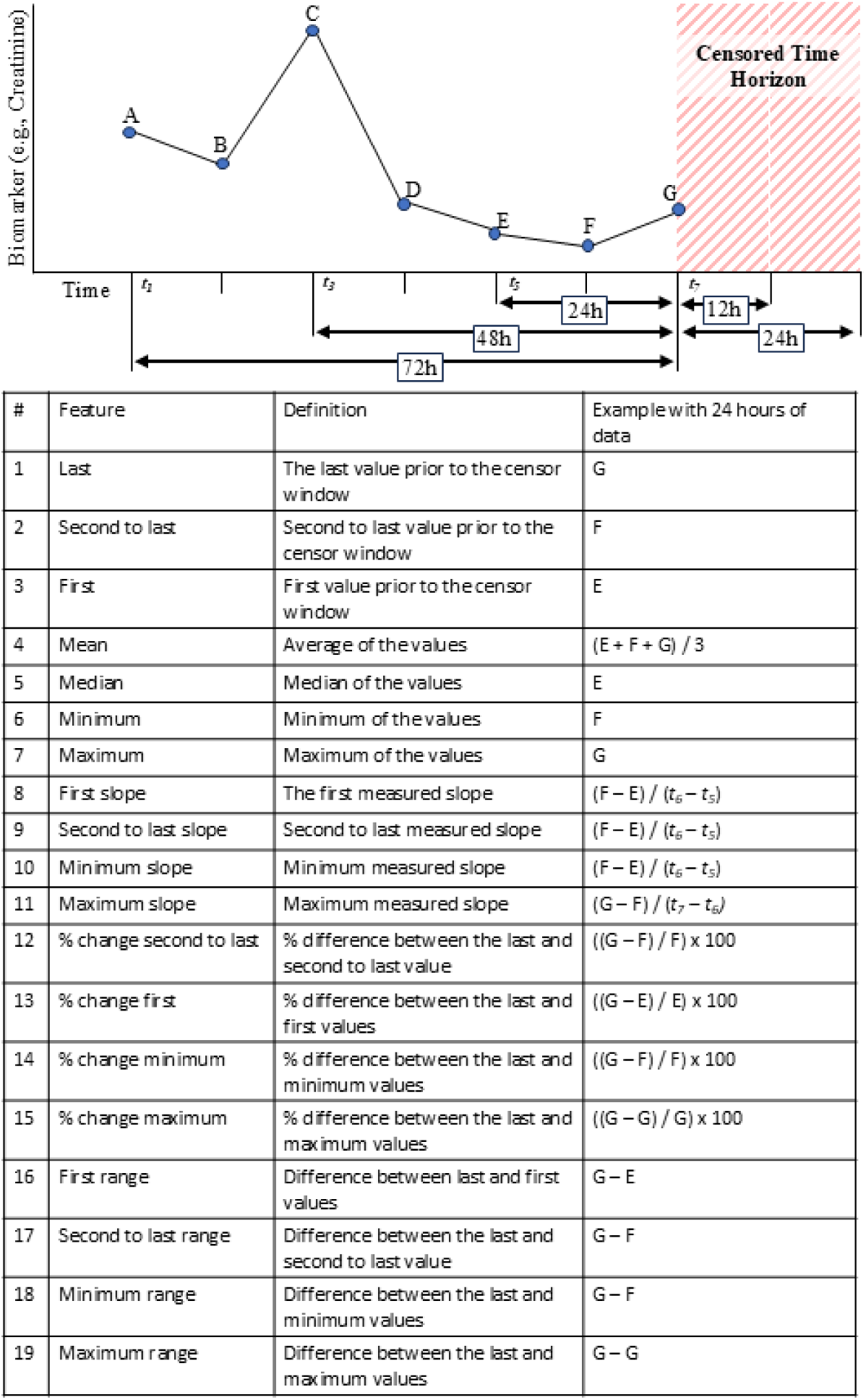
A representation of the feature engineering for continuous biomarker measurements. Temporal information is represented as a variety of summary measurements for discrete windows of time. Feature windows of 24-hours, 48-hours, and 72-hours are displayed in the figure. The models at the development site were trained with 6-hour, 12-hour, and 24-hour censored time horizons, with 12-hour and 24-hour horizons demonstrated in the figure. Definitions and related examples for a 24-hour window of data prior to the censor period are displayed in the table. Abbreviations: h, hours.

**Supplemental Figure 4.**
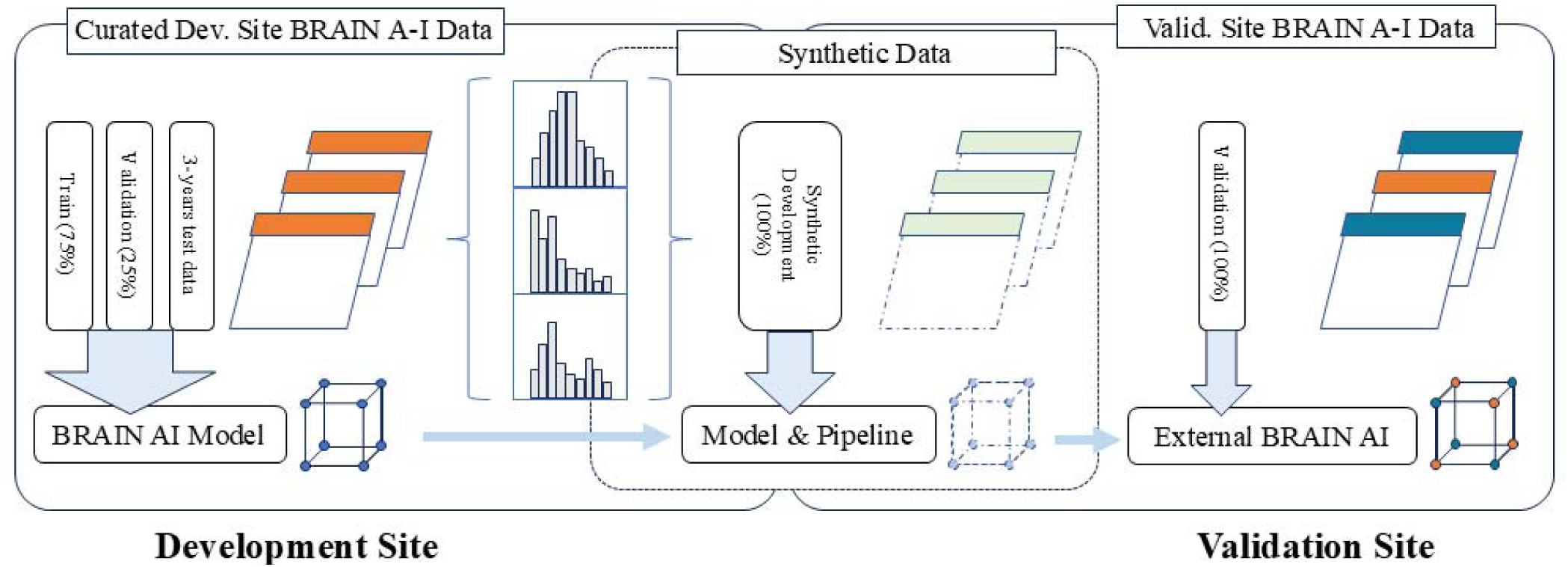
The process of BRAIN A-I model development and external validation. Curated data at the development site were divided into a train cohort, validation cohort, and 2-years of holdout test data. The curated data were used to generate synthetic data with comparable single variable statistical distributions. The synthetic data were then distributed to the external validation site with model training code, facilitating local data curation by providing the necessary details of data structure. Finally, the working BRAIN A-I pipeline was applied to real-world data at the external validation site, applied separate training and validation datasets. Abbreviations: BRAIN A-I, Biodigital Rapid Alert to Identify Neuromorbidity A-I Bundle; Dev. Site, Development Site; Valid. Site, Validation Site.

**Supplemental Figure 5.**
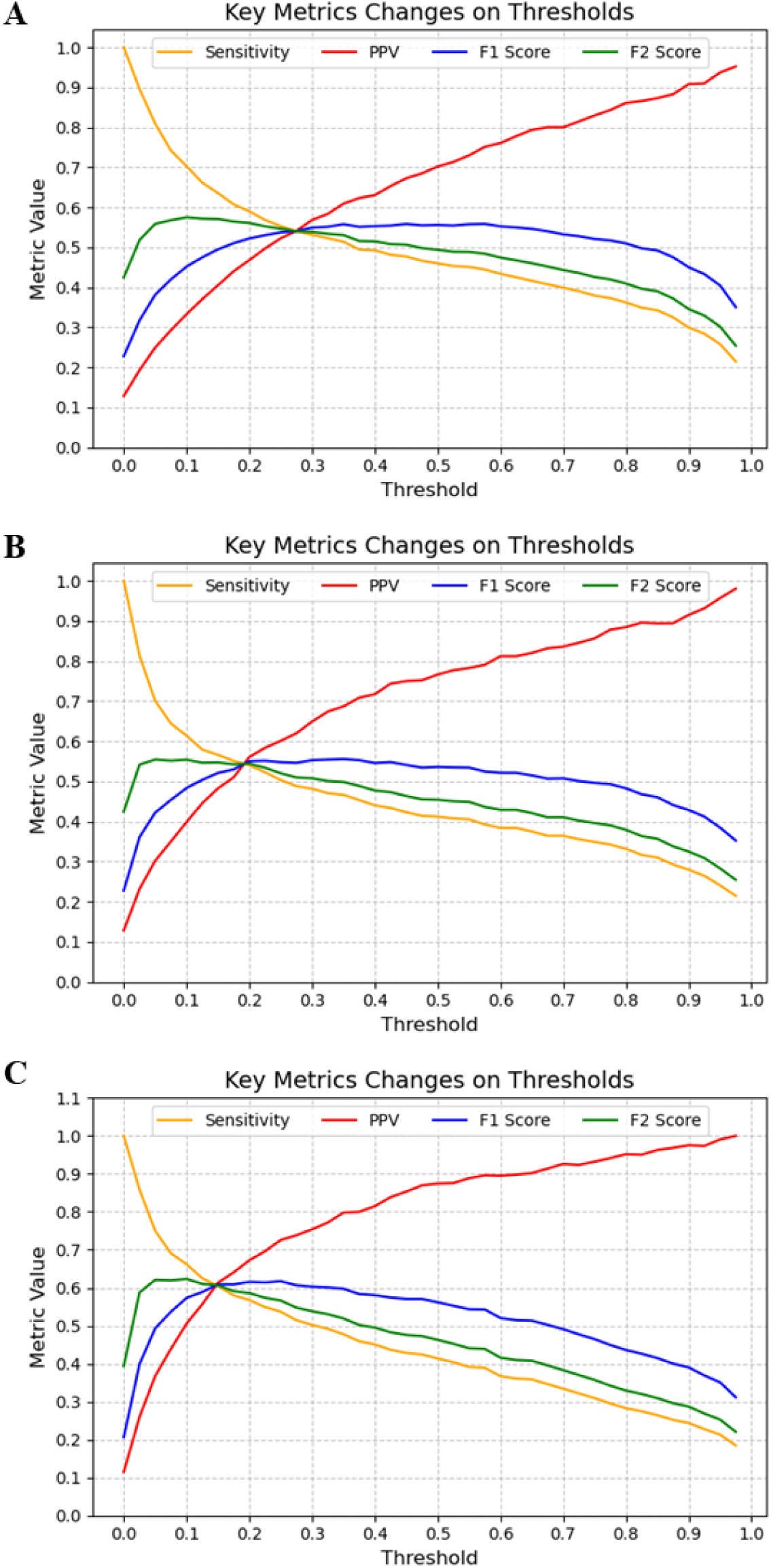
Plots the key statistical performance metrics sensitivity (gold line), positive predictive value (PPV, red line), F1 score (blue line), and F2 score (green line) with metric values on the y-axis and model output thresholds on the x-axis for A) the logistic regression model with a 12-hour time horizon and 48-hour feature window in the development site validation dataset; B) the extreme gradient boosting model with a 12-hour time horizon and 48-hour feature window in the development site validation dataset; and C) the extreme gradient boosting model with a 12-hour time horizon and 48-hour feature window in the development site test dataset.

**Supplemental Figure 6.**
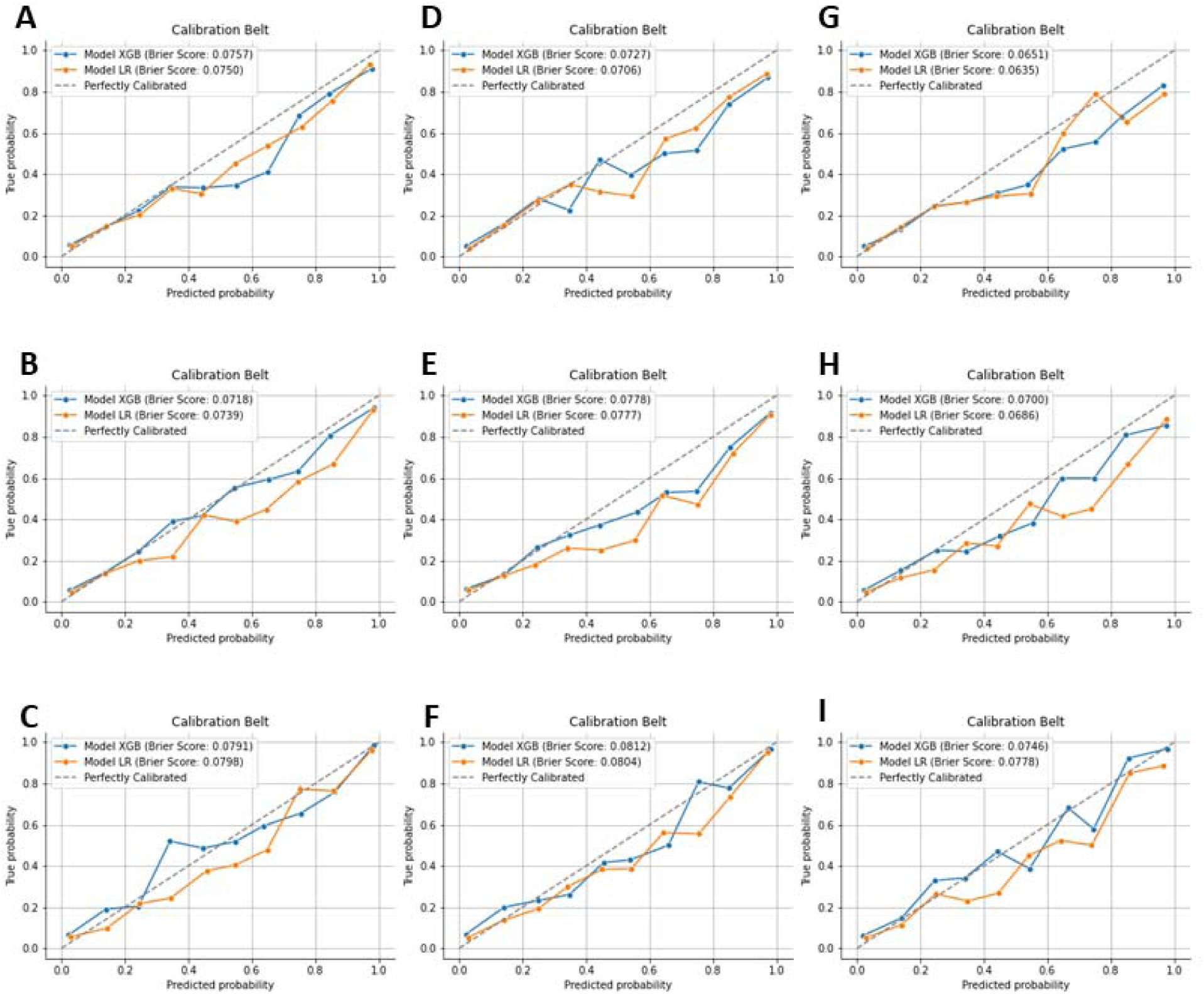
Calibration plots and associated Brier scores for the top performing models for varied time horizons and features windows in the validation dataset. A) 6-hour time horizon and 24-hour feature window; B) 6-hour time horizon and 48-hour feature window; C) 6-hour time horizon and 72-hour feature window; D) 12-hour time horizon and 24-hour feature window; E) 12-hour time horizon and 48-hour feature window; F) 12-hour time horizon and 72-hour feature window; G) 24-hour time horizon and 24-hour feature window; H) 24-hour time horizon and 48-hour feature window; I) 24-hour time horizon and 72-hour feature window. Abbreviations: LR, logistic regression; XGB, extreme gradient boosted.

**Supplemental Figure 7.**
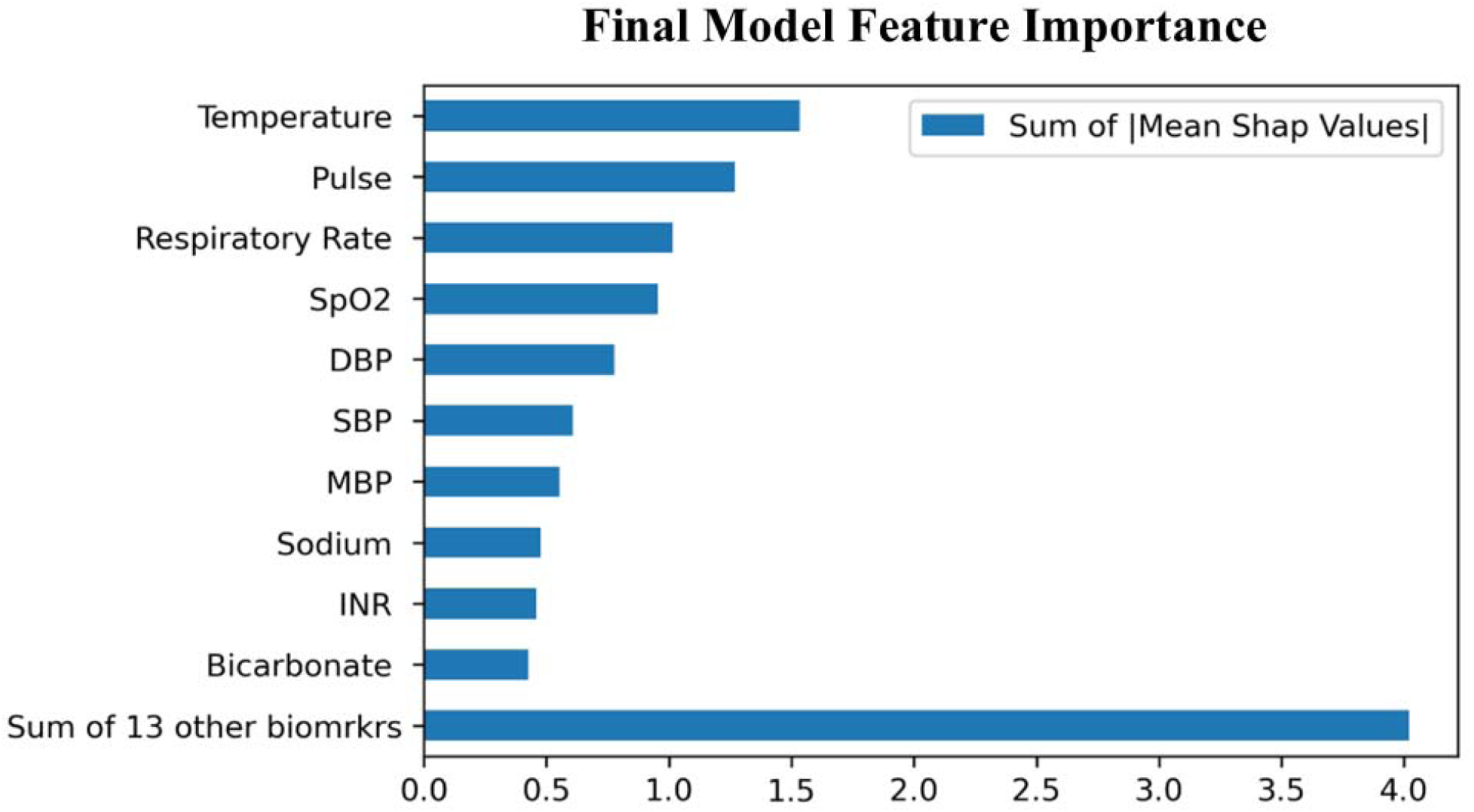
Top 10 biomarker feature categories based on Shap values for the 12-hour time horizon 48-hour feature window XGBoost model from the development site. Each category contains several features, e.g. Temperature contains maximum temperature, minimum temperature, average temperature, etc. The blue bars represent the sum of the mean Shap values for each category. Abbreviations: biomrkrs, biomarkers.

**Supplemental Figure 8.**
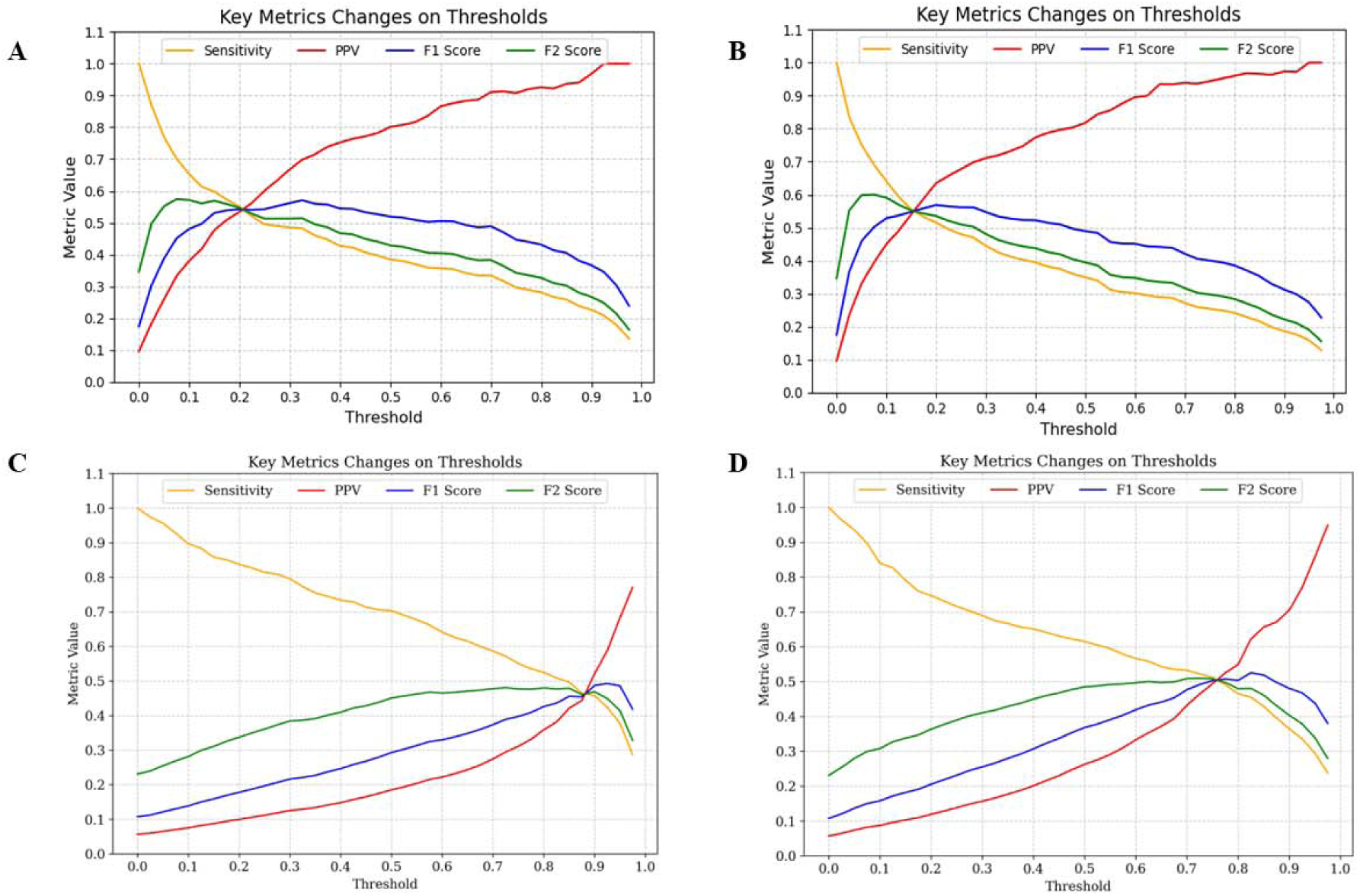
Plots the key statistical performance metrics sensitivity (gold line), positive predictive value (PPV, red line), F1 score (blue line), and F2 score (green line) with metric values on the y-axis and model output thresholds on the x-axis for the generalizable A) logistic regression model in the development site test dataset; B) extreme gradient boosting model in the development site test data set; C) logistic regression model in the external validation dataset; D) extreme gradient boosting model in the external validation site dataset.

**Supplemental Figure 9.**
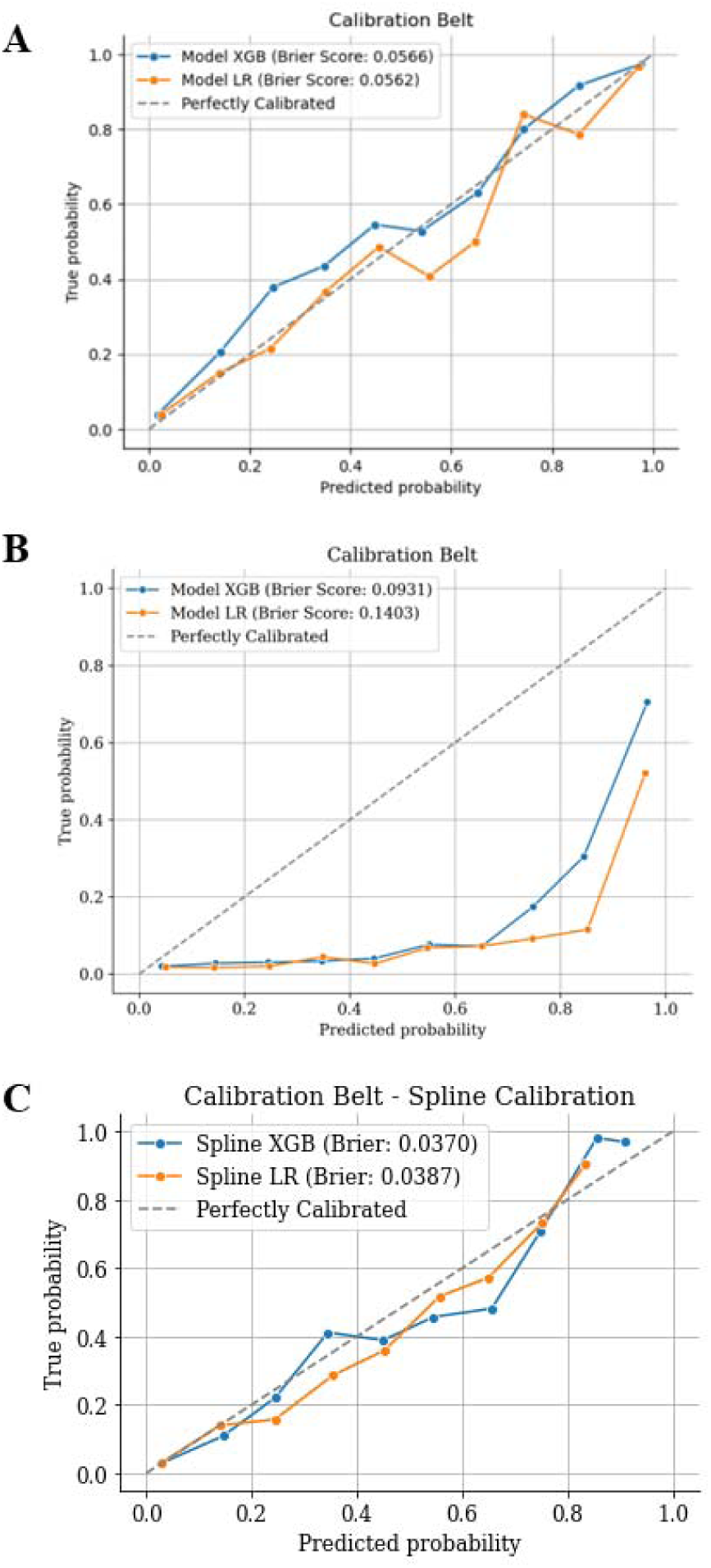
Calibration plots for the generalizable model in the **A**) development site test dataset and **B**) the external validation site dataset.

**Supplemental Figure 10.**
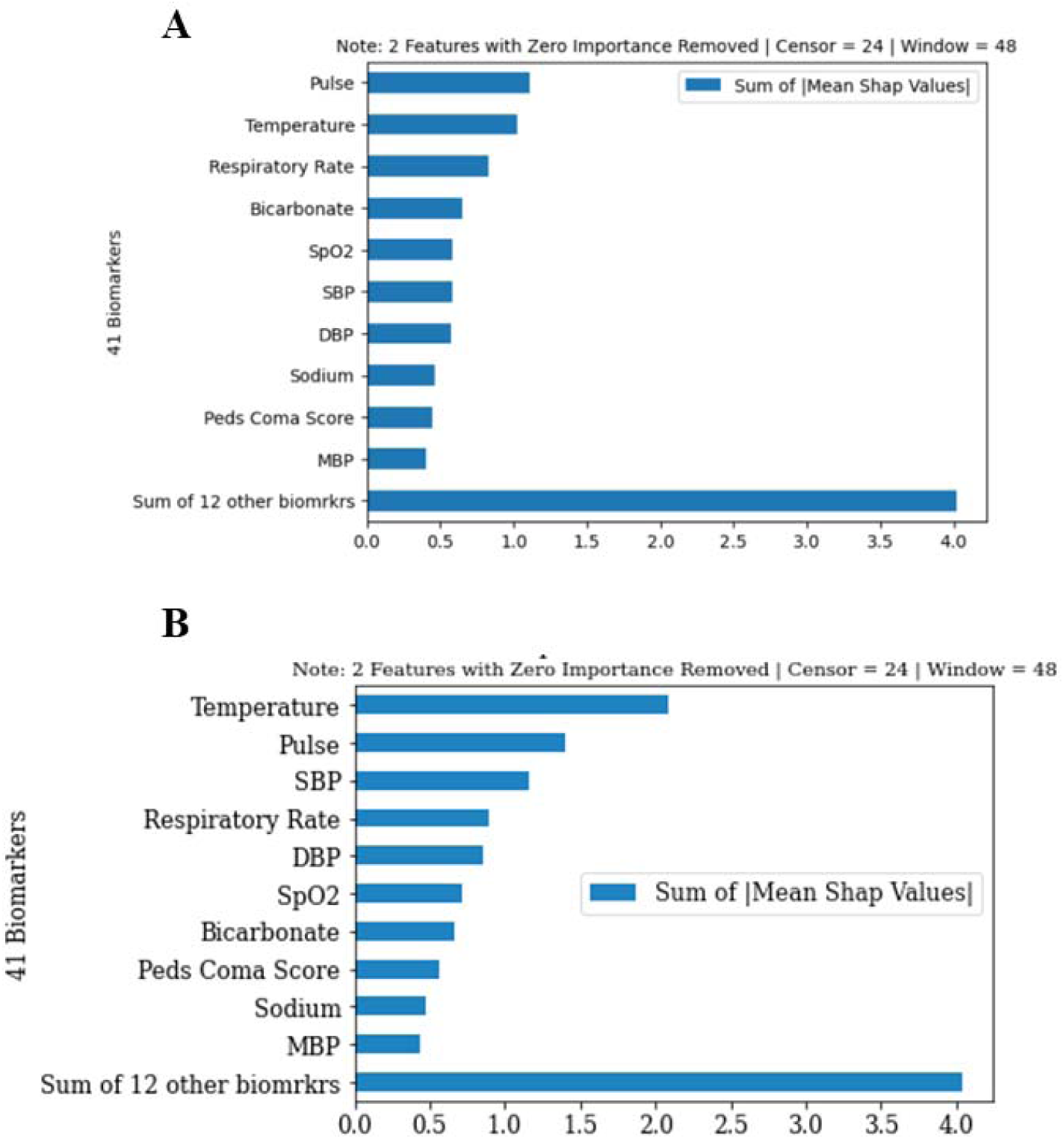
Feature importance analysis for the generalizable model in the **A**) development site test dataset and **B**) external validation site dataset.

## Notes

### Competing Interest Statement

The authors have declared no competing interest.

### Funding Statement

This study was funded by: NINDS R01NS118716 and NLM 5T15LM007059-38.

### Author Declarations

Institutional Review Board of the University of Pittsburgh name gave ethical approval for this work

