## Supplemental Biomarker Methods for "Development, External Validation, and Biomolecular Corroboration of Interoperable Models for Identifying Critically Ill Children at Risk of Neurologic Morbidity"

Blood was obtained by a bedside nurse and set aside in collection tubes, which were then centrifuged at 4^o^ Celsius and spun at 1500xg for 8 minutes. Serum was then retrieved, aliquoted and placed in storage at -80^o^ Celsius. Enzyme-linked immunosorbent assays were used to measure serum levels of neuron specific enolase (NSE), myelin basic protein (MBP), and S100 calcium binding protein B, as previously described.^1,2^ Hemolysis can interfere with measurement of NSE so a correction was applied using a validated formula.^3^ Commercial assays for glial fibrillary acidic protein (GFAP), ubiquitin C-terminal hydrolase (UCH-L1), and alpha-II spectrin breakdown product 150 (SBDP150) were run by Banyan Biomarkers, Inc (San Diego, CA).
